# Prevalent and persistent new-onset autoantibodies in mild to severe COVID-19

**DOI:** 10.1101/2024.02.15.24302857

**Authors:** August Jernbom Falk, Lovisa Skoglund, Elisa Pin, Ronald Sjöberg, Hanna Tegel, Sophia Hober, Elham Rostami, Annica Rasmusson, Janet L. Cunningham, Sebastian Havervall, Charlotte Thålin, Anna Månberg, Peter Nilsson

**Affiliations:** Division of Affinity Proteomics, Department of Protein Science, SciLifeLab, KTH Royal Institute of Technology, Stockholm, Sweden; Division of Protein Technology, Department of Protein Science, KTH Royal Institute of Technology, Stockholm, Sweden; Section of Neurosurgery, Department of Medical Sciences, Uppsala University Hospital, Uppsala, Sweden; Department of Neuroscience, Karolinska Institutet, Stockholm, Sweden; Department of Medical Sciences, Psychiatry, Uppsala University, Uppsala, Sweden; Department of Clinical Sciences, Karolinska Institutet, Danderyd Hospital, Stockholm, Sweden

## Abstract

Autoantibodies have been shown to be implied in COVID-19 but the emerging autoantibody repertoire remains largely unexplored. We investigated the new-onset autoantibody repertoire in 525 healthcare workers and hospitalized COVID-19 patients in five time points over 16 months using proteome-wide and targeted protein and peptide arrays. Our results show that prevalent new-onset autoantibodies against a wide range of antigens emerged following SARS-CoV-2 infection in relation to pre-infectious baseline samples and remained elevated for at least 12 months. We demonstrated associations between distinct new-onset autoantibodies and neuropsychiatric symptoms post-COVID-19. Using epitope mapping, we determined the main epitopes of selected new-onset autoantibodies, validated them in independent cohorts of neuro-COVID and pre-pandemic healthy controls, and identified molecular mimicry between main epitopes and the conserved fusion peptide of the SARS-CoV-2 Spike glycoprotein. Our work describes the complexity and dynamics of the autoantibody repertoire emerging with COVID-19 and supports the need for continued analysis of the new-onset autoantibody repertoire to elucidate the mechanisms of the post-COVID-19 condition.

## Introduction

In SARS-CoV-2 infection^1^ and other pulmonary viral infections^2^, pre-existing anti-type I interferon autoantibodies have been detected in 5-20% of severe disease cases and may affect therapeutic strategies^3,4^. Several studies have detected presence of established autoantibodies in COVID-19 patients^5–10^, although their clinical significance remains unclear. In addition, autoantibodies against a wide range of extracellular antigens have been detected in COVID-19 and a subset of these have been shown to antagonize cytokine signaling, be associated to increased viral loads and decreased T-cell and B-cell populations, and to increase disease severity in mouse models of COVID-19^11^. The total number of these autoantibodies in COVID-19 patients has been associated with disease severity^11^. However, autoantibody repertoires are notoriously individual-specific in both health and disease^12,13^, rendering associations to clinical symptoms and outcomes difficult without a longitudinal study design.

To this end, some studies of hospitalized patients with COVID-19 have investigated development of autoantibodies against a selection of previously described^14^ or extracellular^15^ antigens. While these studies demonstrated the existence of new-onset autoantibodies in patients with severe COVID-19, they were constrained by the use of baseline samples collected after hospitalization and short follow-up times. This presented limitations in evaluation of the persistence of new-onset autoantibodies and their connection to the course of COVID-19.

In the months following COVID-19, an estimated 6% of individuals experience lasting symptoms such as cognitive dysfunction, fatigue, and shortness of breath^16,17^. These symptoms are collectively known as long COVID, post-acute sequelae of COVID-19, or post-COVID-19 condition and may occur after mild as well as severe acute disease^16^. There are many theories on the etiology of the post-COVID-19 condition, including viral persistence, persistent inflammation, and autoimmunity, including emergence of new-onset autoantibodies^3,18–20^. In particular, neurological symptoms after SARS-CoV-2 infection, termed neuro-COVID, are suspected to stem from a dysregulated immune response with autoantibody involvement^5,21–24^, similar to other post-infectious neurological disorders^24,25^.

During the COVID-19 pandemic, we developed a highly specific and sensitive multiplex bead array for SARS-CoV-2 serology^26^ which we have used to profile the serological response in several research projects, where the COMMUNITY (COVID-19 Immunity) study is a longstanding collaboration^27–29^. This ongoing longitudinal study enrolled 2149 healthcare workers (HCW) and 118 admitted COVID-19 patients at Danderyd Hospital, Sweden, between April and May 2020, with follow-up visits every four months.

In the present study, we extend the analysis within a subgroup of the COMMUNITY study cohort by profiling the dynamics of autoantibody repertoires across SARS-CoV-2 infection using proteome-wide and targeted in-house developed planar and bead arrays. The results reveal prevalent new-onset autoantibodies against a wide range of antigens which remain elevated for at least 12 months, are associated to neuropsychiatric symptoms post-COVID-19, and show molecular mimicry with the SARS-CoV-2 Spike protein fusion peptide.

## Results

In this study, we have profiled the autoantibody repertoire of 478 HCW and 47 hospitalized COVID-19 patients (Table S1). An overview of the study is shown in Figure 1. In summary, samples were collected across 3-5 visits (mean 4.8) over 16 months, for a total of 2532 samples analyzed in the present study. In HCW, 20% (n=96) were seropositive (had anti-SARS-CoV-2 immunoglobulin gamma (IgG)) at study inclusion in May 2020. Among the remaining 382 baseline seronegative HCW, 109 seroconverted (first display of anti-SARS-CoV-2 IgG) in Sept 2020, 233 in Jan 2021, and 40 in May 2021. All HCW seroconverted before receiving their first SARS-CoV-2 vaccine dose. Among the patients, 85% (n=40) were seropositive at the first sampling after admission (May 2020), and the remaining 15% (n=7) had seroconverted at the first sampling after discharge (Sept 2020).

**Fig. 1.**
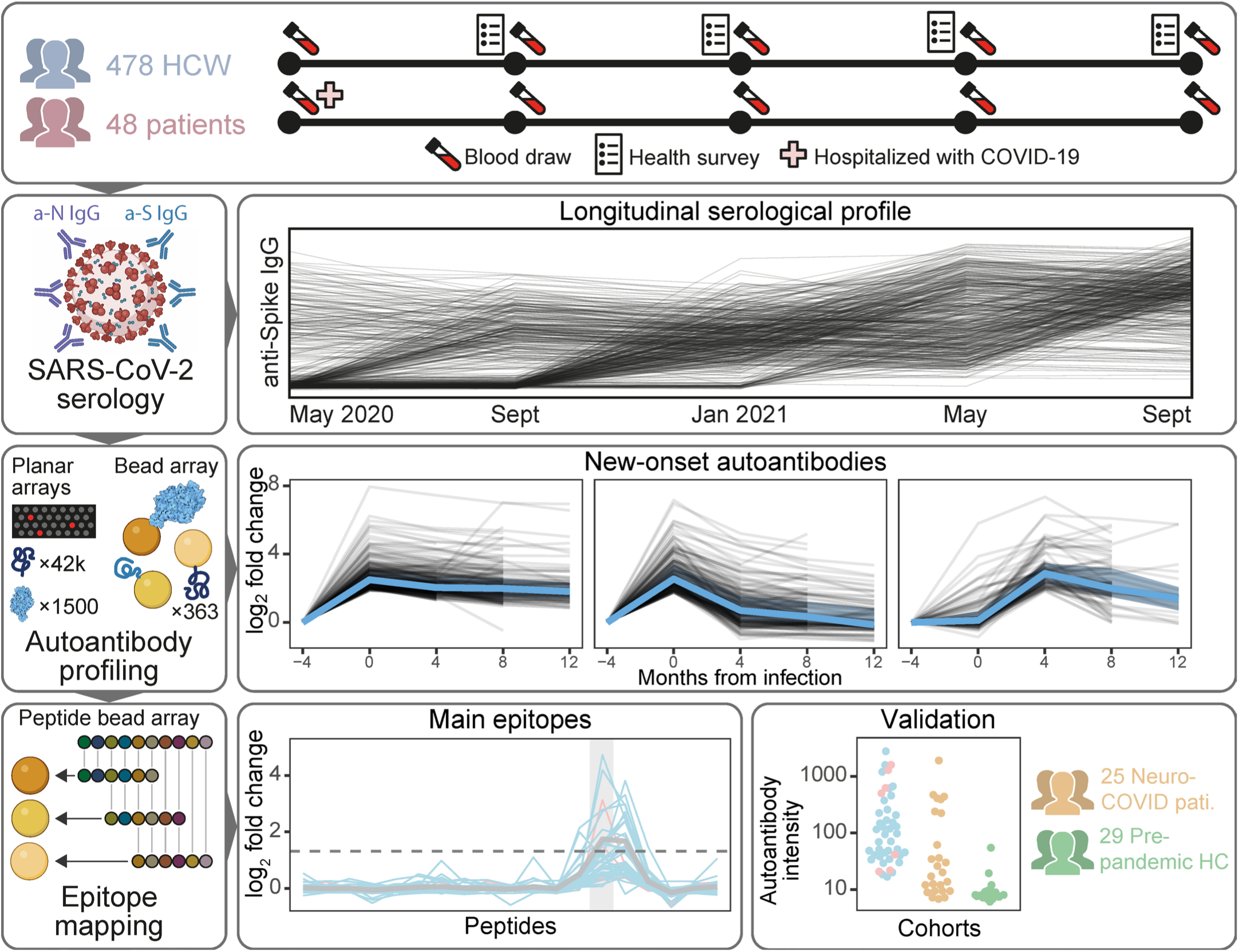
Study overview. Healthcare workers (HCW, n=478) and patients (n=48) with complete sample sets from May 2020 to Sept 2021 were selected for the study. Longitudinal serological profiles were obtained using our in-house SARS-CoV-2 serology assay. Proteome-wide planar and bead arrays were used to chart the autoantibody repertoire across seroconversion for identification of new-onset autoantibodies. Tiled peptide bead arrays were used to identify main epitopes of selected new-onset autoantibodies and validate them in independent cohorts.

### Proteome-wide autoantibody profiling reveals diverse autoantibodies in COVID-19

To explore the proteome-wide autoantibody landscape emerging with COVID-19, we screened blood sample from 32 healthcare workers (HCW) with self-reported symptoms post-COVID-19, and 16 hospitalized COVID-19 patients, on our in-house developed planar array platforms. Plasma samples from the 32 HCW were divided in eight groups of four individuals each, defined by specific symptoms post-COVID-19: anosmia or ageusia; palpitations and shortness of breath; cognitive and neurological symptoms; dermatological symptoms; fever, fatigue, and bodily pain; multiple and severe symptoms; new diagnoses after COVID-19; multiple symptoms after severe COVID-19. Similarly, plasma samples from the 16 patients were divided in four groups of four individuals each, based on sex and comorbidities: male patients 1, male patients 2, female patients, and patients with comorbidities. Within each group, plasma samples were combined and analyzed on the arrays. HCW and patient groups were analyzed on the Proteome-wide arrays containing 42 000 protein fragments, and HCW groups were in addition analyzed on the Secretome arrays containing 1522 full-length proteins.

In total, IgG binding was detected towards 215 protein fragments and 22 full-length proteins, with 14 to 36 reactive autoantibodies in each group. Autoantibody profiles were highly specific to each set of combined samples, with 6% (15 of 237) of autoantibodies being reactive in two groups and 3% (8 of 237) being reactive in three to six groups (Fig. S1). The reactive antigens were selected for further investigation in the full HCW and patient cohorts. In addition, antigens were selected by combining evidence from multiple groups and arrays with prior knowledge from literature and in-house studies. The final panel included 307 protein fragments and 56 full-length proteins. Together, these 363 antigens represented proteins from 315 genes (Table S2).

### Prevalent and persistent new-onset autoantibodies emerge with COVID-19

For initial investigation of new-onset autoantibodies, data from the full HCW and patient cohorts were filtered to include individuals with three consecutive samplings before, at, and after seroconversion (n=369: 362 HCW and 7 hospitalized patients). Individual autoantibody trajectories were defined by calculating fold change (FC) of autoantibody levels at the two later time points relative to the seronegative baseline. Clustering of all obtained trajectories revealed three distinct categories of new-onset autoantibodies shown in Figure 2a: stable (n=225), transient (n=177), and delayed (n=103) new-onset autoantibodies. Four additional clusters of relatively unchanging trajectories were detected and not classified as new-onset (Fig. S2). The new-onset autoantibody landscape of individuals and antigens with at least one detected new-onset autoantibody (n individuals = 204, n antigens = 187) is displayed in Figure S3. These autoantigens represent extracellular (n=57) and intracellular (n=119) proteins corresponding to 176 genes as classified in the Human Protein Atlas^30^.

**Fig. 2.**
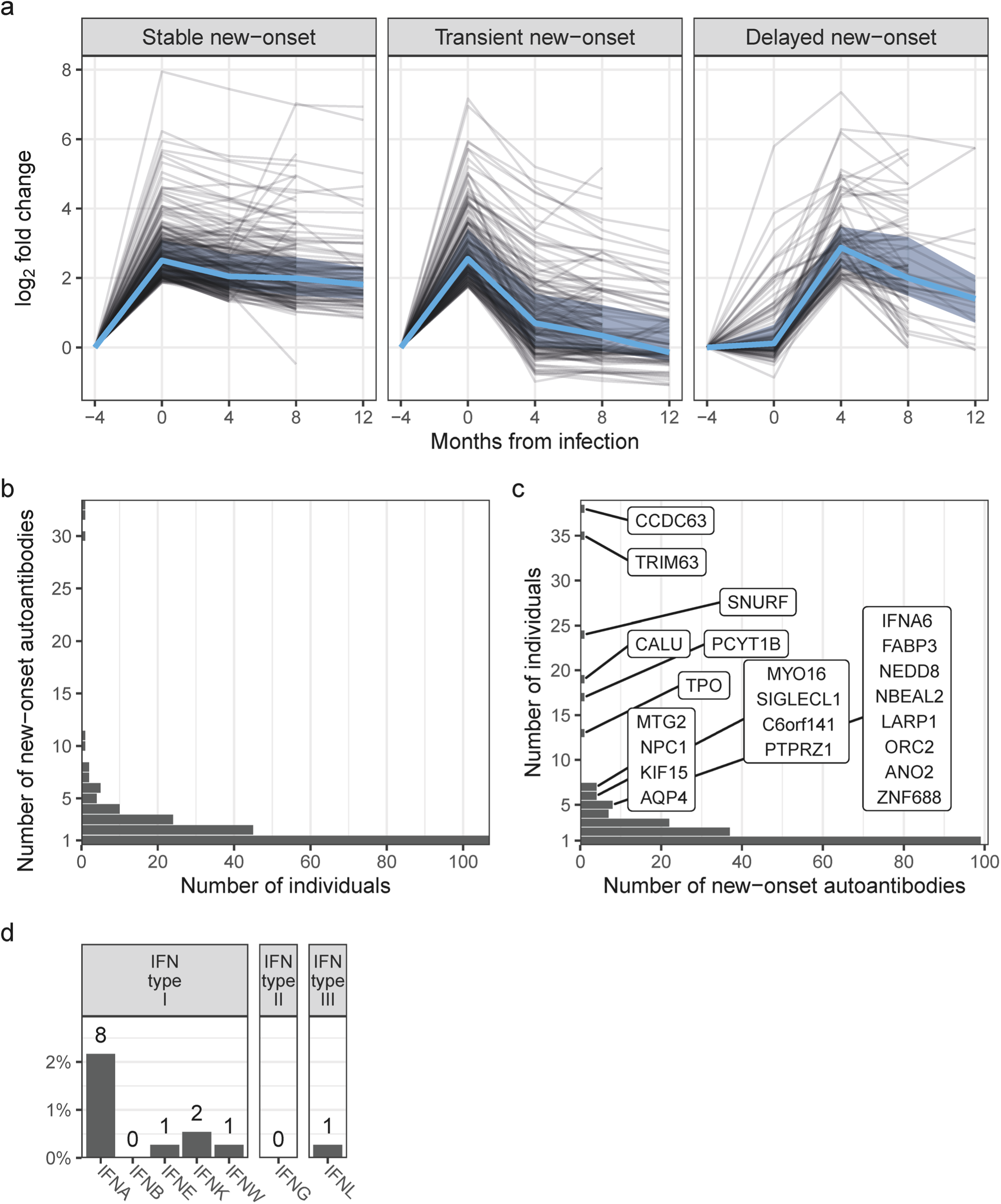
Prevalent and persistent new-onset autoantibodies emerge with COVID-19 and are independent of anti-Spike IgG levels. **a** Persistence of new-onset autoantibodies across categories. New-onset autoantibodies were categorized by their dynamics: Stable, transient, or delayed. Black lines represent new-onset autoantibody trajectories based on fold change in relation to seronegative baseline. Blue lines and shaded areas represent median and quartiles, respectively. **b** Distribution of new-onset autoantibodies among baseline seronegative individuals. Bars depict the number of individuals with the indicated number of new-onset autoantibodies. **c** Distribution of new-onset autoantibody prevalence. Bars depict the number of new-onset autoantibodies with the indicated prevalence. The 22 prevalent new-onset autoantibodies with prevalence >1% (number of individuals >4) are indicated with gene names. **d** Prevalence of new-onset autoantibodies towards IFN subtypes.

We further examined the persistence of new-onset autoantibodies in the 160 trajectories where 12-month follow-up data was available (stable and transient trajectories in 63 individuals). Persistence, defined as FC ≥ 2 at 12 months compared to baseline, was observed for the majority of autoantibodies with stable trajectories (95% (78/82)), while only 23% (18/78) of transient new-onset autoantibodies remained elevated. In total, 60% of new-onset autoantibodies remained elevated 12 months after onset.

Further investigation of the trajectories revealed that new-onset autoantibodies were found in 204 of the 369 individuals and that they targeted in total 187 antigens. Most individuals displayed single new-onset autoantibodies (n=107) but three individuals we found to have 30 to 33 (Fig. 2b). In line with previous reports^11,13,31^, most autoantibodies (99 of 187 detected) were rare and occurred in single individuals (Fig. 2c). The 22 most prevalent new-onset autoantibodies, detected in >1% of the cohort (>4 individuals), were subjected to further analysis (Table 1). The corresponding antigens represented both intracellular (n=16, 73%) and extracellular (n=6, 27%) proteins. Autoantibodies which have previously been reported in autoimmune diseases or COVID-19 patients were present among the most prevalent new-onset autoantibodies, including anti-TPO (thyroid peroxidase)^10^, anti-AQP4 (aquaporin 4)^9^ and anti-IFNA^1^ IgG. However, emergence of these autoantibodies with COVID-19 has not been reported previously. Several of the most prevalent new-onset autoantibodies have to our knowledge not been described previously, including the three with the highest prevalence, i.e., anti-CCDC63 (coiled-coil domain-containing protein 63), anti-TRIM63 (E3 ubiquitin-protein ligase TRIM63), and anti-SNURF (SNRPN upstream reading frame protein) IgG. In total, 150 of the 369 baseline seronegative individuals (41%) developed at least one of the 22 most prevalent new-onset autoantibodies.

**Table 1.** Autoantibodies with >1% prevalence.

| Gene | Protein | Antigen | ENSP | aa | N new-onset | Acute onset (%) | Baseline seronegative (%) |
| --- | --- | --- | --- | --- | --- | --- | --- |
| CCDC63 | Coiled-coil domain-containing protein 63 | HPRR3070734 | ENSP00000312399 | 155-249 | 49 | 96 | 78 |
| SNURF | SNRPN upstream reading frame protein | HPRR4280292 | ENSP00000463201 | 45-69 | 49 | 76 | 49 |
| TRIM63 | E3 ubiquitin-protein ligase TRIM63 | HPRR3760711 | ENSP00000363390 | 182-246 | 44 | 66 | 80 |
| CALU | Calumenin | HPRR4180820 | ENSP00000249364 | 74-128 | 32 | 41 | 59 |
| TPO | Thyroid peroxidase | HPRR4180385 | ENSP00000329869 | 867-932 | 22 | 95 | 59 |
| PCYT1B | Choline-phosphate cytidyltransferase B | HPRR4320357 | ENSP00000368439 | 329-369 | 22 | 86 | 77 |
| MYO16 | Unconventional myosin-XVI | HPRR3090005 | ENSP00000401633 | 843-923 | 18 | 89 | 33 |
| AQP4 | Aquaporin-4 | HPRR2140210 | ENSP00000372654 | 253-323 | 17 | 35 | 41 |
| NPC1 | NPC intracellular cholesterol transporter 1 | HPRR620021 | ENSP00000269228 | 448-580 | 14 | 79 | 50 |
| NBEAL2 | Neurobeachin-like protein 2 | HPRR3460329 | ENSP00000415034 | 1865-1939 | 13 | 100 | 38 |
| ZNF688 | Zinc finger protein 688 | HPRR3610292 | ENSP00000223459 | 95-166 | 12 | 92 | 42 |
| IFNA6 | Interferon alpha-6 | HPRR3360040 | ENSP00000369558 | 74-99 | 12 | 25 | 42 |
| SIGLECL1 | SIGLEC family-like protein 1 | HPRR3420383 | ENSP00000469601 | 11-94 | 10 | 90 | 60 |
| MTG2 | Mitochondrial ribosome-associated GTPase 2 | HPRR3300084 | ENSP00000359859 | 317-390 | 10 | 80 | 70 |
| NEDD8 | NEDD8 | HPRR2760190 | ENSP00000250495 | 7-67 | 10 | 70 | 50 |
| KIF15 | Kinesin-like protein KIF15 | HPRR2960531 | ENSP00000324020 | 671-748 | 10 | 60 | 70 |
| FABP3 | Fatty acid-binding protein, heart | HPRR3890315 | ENSP00000362817 | 30-58 | 9 | 67 | 56 |
| ANO2 | Anoctamin-2 | HPRR3070036 | ENSP00000507275 | 79-167 | 9 | 56 | 56 |
| LARP1 | La-related protein 1 | HPRR3760556 | ENSP00000428589 | 254-329 | 9 | 44 | 56 |
| ORC2 | Origin recognition complex subunit 2 | HPRR2920267 | ENSP00000234296 | 150-224 | 8 | 75 | 62 |
| C6orf141 | Uncharacterized protein C6orf141 | HPRR2660035 | ENSP00000434602 | 5-73 | 7 | 86 | 86 |
| PTPRZ1 | Receptor-type tyrosine-protein phosphatase zeta | HPRR4170025 | ENSP00000377047 | 441-534 | 6 | 50 | 100 |

Considering the demonstrated impact of antibodies targeting interferons (IFNs) in COVID-19^1^, we specifically investigated the prevalence of new-onset autoantibodies across IFN subtypes. In total, we detected new-onset anti-IFN IgG in 10 of 362 baseline seronegative HCW and 1 of the 7 COVID-19 patients that were seronegative at admission. Anti-interferon alpha (IFNA) IgG was predominant (Fig. 2d), and 1 individual had new-onset autoantibodies targeting more than one IFN subtype (IFNA, interferon epsilon (IFNE), and interferon omega (IFNW)).

### Prevalent new-onset autoantibodies in individuals without pre-infectious samples

Next, we aimed to explore associations of the 22 most prevalent new-onset autoantibodies to COVID-19 severity and symptoms post-COVID-19. To include individuals who were seropositive at study inclusion (94 HCW and 39 hospitalized patients), we first developed a model for classification of new-onset based on autoantibody levels at seroconversion and four- and eight-month follow-up. The model used the aggregated categories acute new onset (stable or transient) and delayed new-onset.

This multinomial linear regression (MNL) model was trained on the autoantibody trajectories of the previously assessed baseline seronegative individuals that had an eight-month follow-up sample after seroconversion (n individuals = 282; n trajectories for training = 6204). Using an 80%/20% training/testing split, we found that the model performed well on the test set and was highly specific although moderately sensitive (specificity = 0.997, sensitivity = 0.667, AUC = 0.83). Applied to the baseline seropositive individuals, the model classified 98 autoantibody trajectories as acute new-onset and 56 as delayed new-onset in 79 individuals (59%) (Fig. S4). The high specificity and moderate sensitivity of the model indicates that it may underestimate new-onset autoantibody prevalence in individuals without baseline seronegative samples. Still, the 22 new-onset autoantibodies were significantly more prevalent in individuals without baseline seronegative samples than in individuals with baseline seronegative samples (average prevalence 5.3% and 2.9%, respectively; odds ratio (OR) = 1.42, p = 4×10^-6^, n trajectories = 11044). This could indicate increased autoimmune signatures in the hospitalized COVID-19 patients.

Summarizing the new-onset autoantibody landscape in both seronegative and seropositive baseline individuals, 43% of HCW (n=196/456) and 72% of hospitalized COVID-19 patients (n=33/46) displayed at least one of the most prevalent new-onset autoantibodies (Fig. 3a). As shown in Figure 3b, 10 of the 22 were significantly more prevalent in the patients than the HCW, regardless of the number of symptoms post-COVID-19 (logistic regression model adjusted for age and sex, Benjamini-Hochberg correction, q ≤ 0.05).

**Fig. 3.**
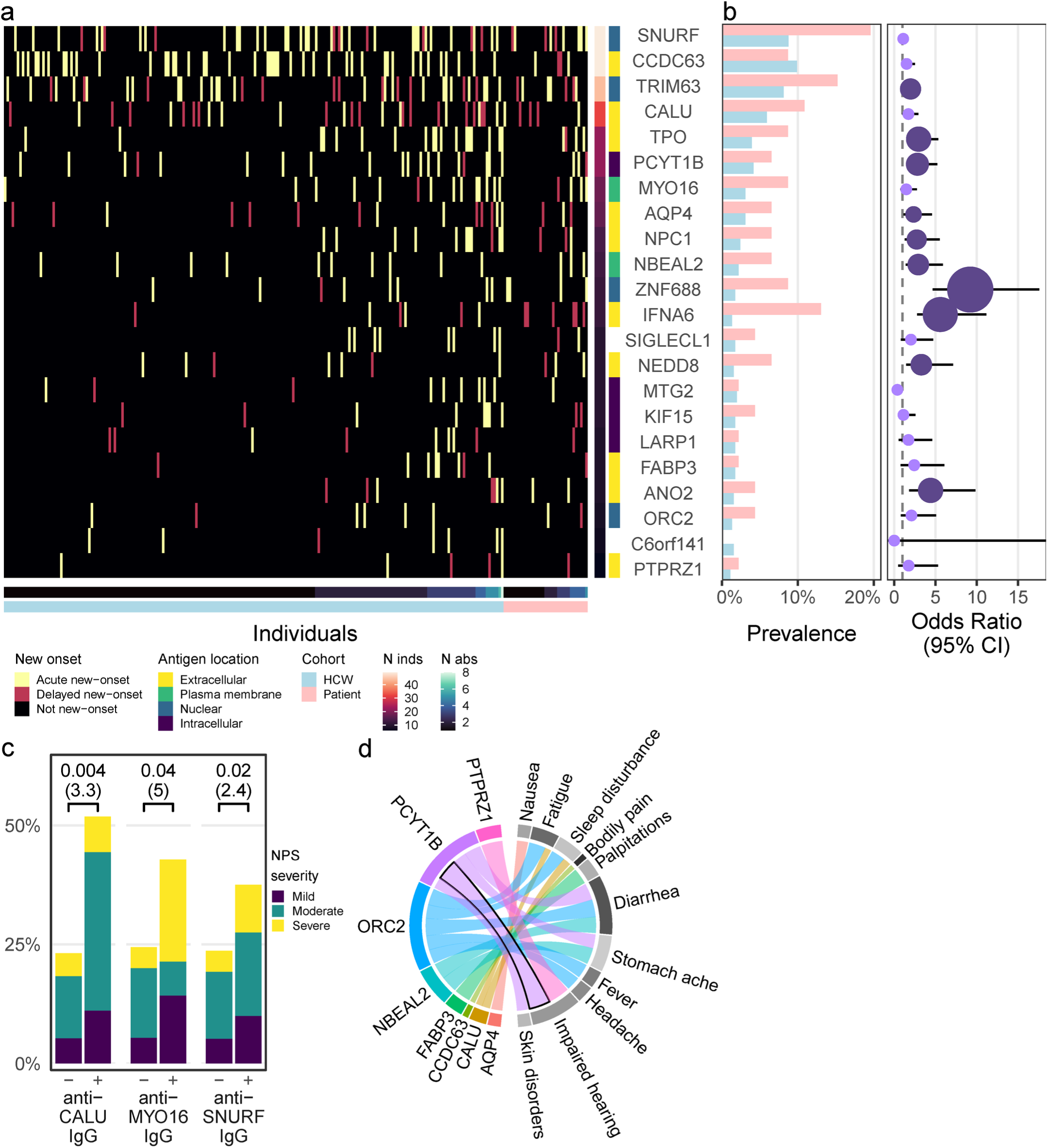
New-onset autoantibodies have increased prevalence in hospitalized COVID-19 patients and are associated with neuropsychiatric symptoms post-COVID-19. **a** Overview of new-onset autoantibodies in HCW and hospitalized COVID-19 patients. The 22 prevalent new-onset autoantibodies are grouped by their antigen location. Columns show individuals with at least one prevalent new-onset autoantibody. Cell color represents new-onset autoantibody category: Acute, delayed, or not new-onset. **b** Hospitalized COVID-19 patients showed increased prevalence of nearly half of the most prevalent new-onset autoantibodies. Bars indicate autoantibody prevalence in patients (pink) and HCW (blue). Point position and lines indicate odds ratio with 95% confidence interval (CI) based on logistic regression. Point size and color shows magnitude of q-value (Benjamini-Hochberg). Light purple: q > 0.05. Dark purple: q ≤ 0.05. **c** Increased severity of neuropsychiatric symptoms post-COVID-19 was associated with three new-onset autoantibodies. Bars depict proportions of HCW with different severities of neuropsychiatric symptoms post-COVID-19 among autoantibody-positive (+) and negative (-) groups. Brackets indicate p-values and odds ratios (in parentheses) from proportional odds logistic regression. **d** Eleven symptoms post-COVID-19 were associated with eight of the most prevalent new-onset autoantibodies. Band widths indicate the estimated logit (log odds ratio) of associations with p ≤ 0.05. The indicated association of PCYT1B and impaired hearing remained significant after Benjamini-Hochberg correction (q = 0.002).

New-onset autoantibodies are associated with neuropsychiatric symptoms post-COVID-19 Next, we investigated whether the most prevalent new-onset autoantibodies were associated with neuropsychiatric symptoms post-COVID-19 in HCW. Using a proportional odds logistic regression model, we identified three new-onset autoantibodies associated with increased odds of higher severity of neuropsychiatric symptoms lasting for at least 2 months post-COVID-19, shown in Figure 3c; anti-CALU (calumenin), anti-MYO16 (unconventional myosin-XVI), and anti-SNURF IgG (OR = 3.3, 5.0, 2.4; p = 0.004, 0.04, 0.02, respectively). In addition, we asked whether the most prevalent new-onset autoantibodies were associated to other post-COVID-19 symptoms. Using logistic regression, we identified 8 autoantibodies associated to 11 other reported moderate or severe symptoms post-COVID-19 with the association between anti-PCYT1B (choline-phosphate cytidylyltransferase B) IgG and impaired hearing being the strongest (OR=41, q=0.002; Fig. 3d). Interestingly, there was also a moderate association between autoantibodies towards the muscle protein CCDC63 and muscle and joint pain (OR=2.3, p=0.03), although this association was not significant after Benjamini-Hochberg correction.

### Anti-SNURF IgG increases at vaccination

While all 456 HCW that were assessed for new-onset autoantibodies had seroconverted before vaccination, 362 (79%) received their first SARS-CoV-2 vaccine dose during the study period. The majority received the Pfizer/BioNTech vaccine (68%, n=248), 30% received the AstraZeneca vaccine (n=107), and 2% received the Moderna vaccine (n=7). With this in mind, we asked whether any of the 22 most prevalent autoantibodies increase after vaccination. Considering changes at the level of log_2_ FC ≥ 2, we identified 12 autoantibodies in 45 individuals (Fig. S5). Notably, anti-SNURF IgG was the most commonly increasing autoantibody after vaccination (67%; detected in 30 of 45 HCW displaying an increase). As seen in Figure 4, individuals with as well as without previous new-onset anti-SNURF IgG displayed increases in anti-SNURF IgG at vaccination (Fig. 4b) at comparable levels that of new-onset (Fig. 4a). However, the odds were greater for HCW with previous new-onset anti-SNURF IgG (5 of 28 vs 25 of 332, respectively; OR=3.4, p=0.03, n=360). As four of these five individuals had received the AstraZeneca SARS-CoV-2 vaccine, we investigated whether the odds of autoantibody increase at vaccination was influenced by any interaction effect of previous new-onset autoantibodies and vaccine type and did not find sufficient evidence to support this notion (OR=10, p=0.08, n=360).

**Fig. 4.**
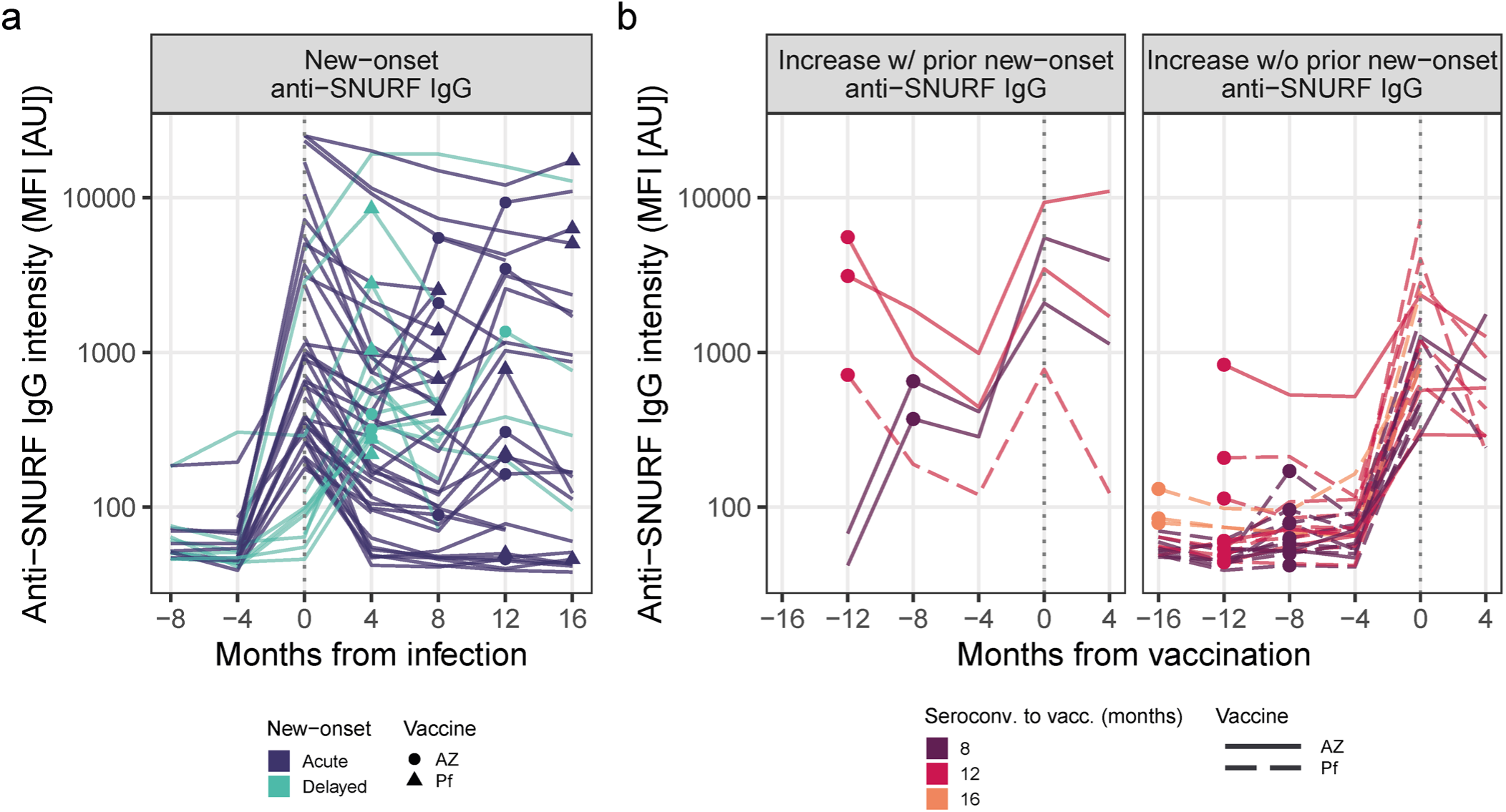
Increasing levels of anti-SNURF IgG at vaccination. Line plots of anti-SNURF IgG dynamics. **a** Individuals with new-onset anti-SNURF IgG. Line color: mode of new-onset. Points: time of vaccination. Point shape: vaccine type. **b** Individuals with 4-fold increase of anti-SNURF IgG levels at vaccination vs 4 months prior. Individuals are separated across panels based on any prior new-onset anti-SNURF IgG at infection. Line color: time between seroconversion and first vaccination. Points: time of seroconversion. Line type: vaccine type.

### Main epitopes of new-onset autoantibodies

Furthermore, we asked what epitopes were targeted by new-onset autoantibodies. To address this question, we epitope mapped eight of the 22 most prevalent new-onset autoantibodies: the highly prevalent anti-CALU, CCDC63, SNURF, and TRIM63 IgG; the previously described anti-IFNA6, ANO2 (anoctamine 2), and TPO IgG; and anti-NPC1 (NPC intracellular cholesterol transporter 1) IgG for which the antigen has sequence overlap with TPO. The epitope mapping was performed on samples from the 142 individuals that had one or more of the eight selected new-onset autoantibodies, using an array of custom designed 14- and 15-mer peptides with an overlap of 10 to 13 amino acid residues (Table S3).

Five autoantibodies displayed main epitopes that were common to individuals with the corresponding new-onset autoantibody. The epitopes correspond to the peptides CCDC63|175-189, NPC1|566-580, SNURF|50-64, TPO|918-932, TRIM63|234-247, TRIM63|236-249, and ANO2|135-149 (not shown).

Aside from the main epitopes, other epitopes occurred individually (Fig. 5a). We next investigated the correspondence of autoantibodies targeting the main epitopes and autoantibodies detected against the full antigen. We observed significantly elevated log_2_ FC of autoantibodies against the main epitopes in individuals with vs without the respective new-onset autoantibody, except in anti-ANO2|135-149 IgG which therefore was excluded from further analysis (Fig. S6). The correlation of autoantibodies detected using different antigen representations varied from a general correlation, e.g., for the SNURF antigens, to a weak correlation in the CCDC63 antigens (Fig. 5c).

**Fig. 5.**
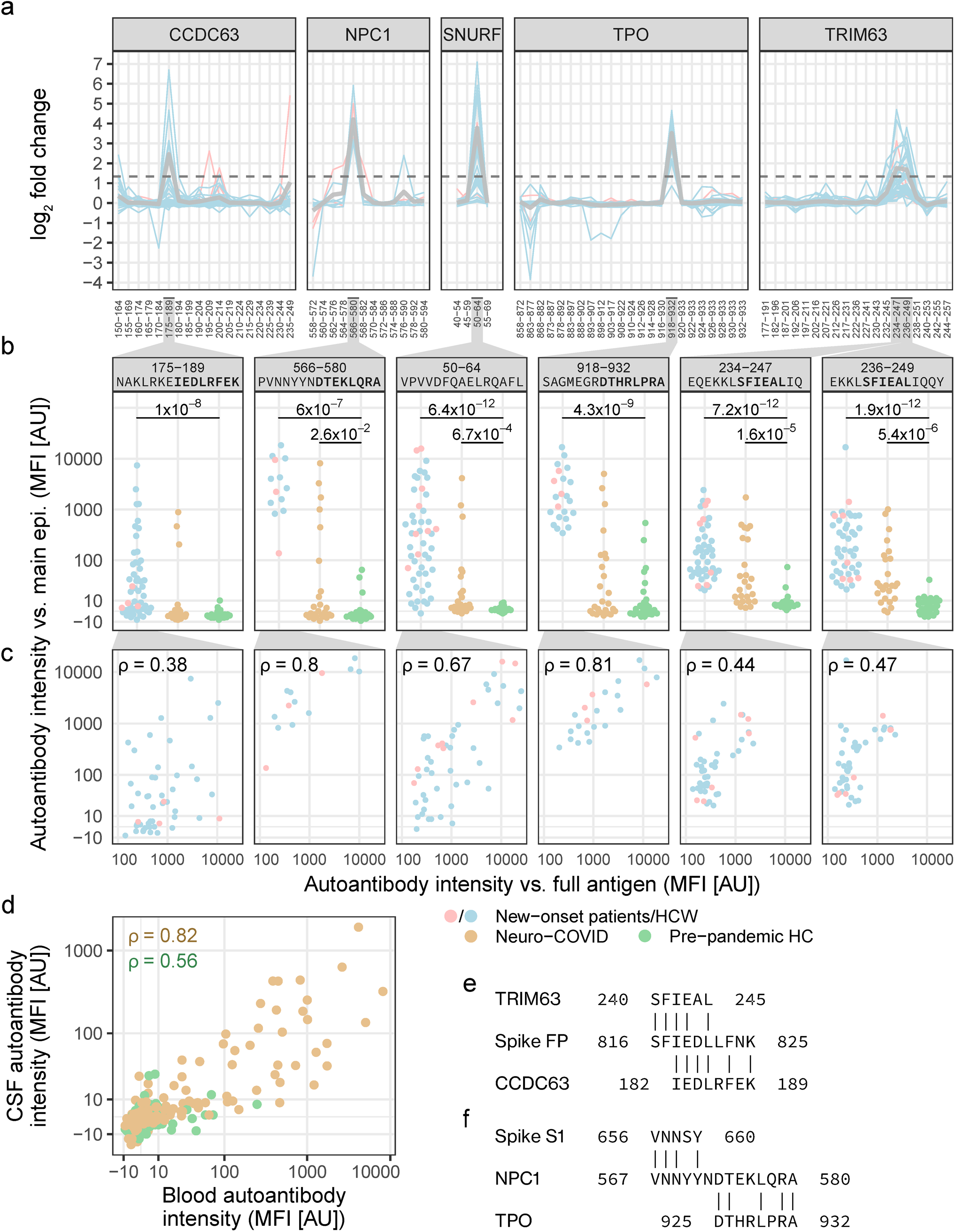
The main epitopes of new-onset autoantibodies are elevated in blood and CSF of an independent cohort with neuro-COVID and display molecular mimicry with the SARS-CoV-2 fusion peptide. **a** Epitope mapping revealed the main epitopes of six new-onset autoantibodies. Lines depict epitope profiles (anti-peptide IgG log_2_ FC) at new-onset in individuals with the corresponding new-onset autoantibody. Gray lines depict the mean. The dashed line indicates the cutoff for classification as a main epitope (FC≥2.52). Numbering on the x-axis corresponds to the peptide amino acid positions in the full-length protein (Table S3). **b** Antibodies against the main epitopes were elevated in an independent cohort of neuro-COVID patients compared to pre-pandemic HCs. Brackets indicate statistically significant difference to pre-pandemic HC (q-values ≤ 0.05 from Mann-Whitney U-test with Benjamini-Hochberg correction). The y-axis displays signal intensity on the pseudo-log_10_ scale. **c** Correlation of autoantibody intensity against the main epitopes (epi., y axis) and the full antigen (x axis). ρ: Spearman’s rho. **d** Correlation of autoantibodies against the main epitopes in blood and CSF of neuro-COVID patients and pre-pandemic HCs. Data points correspond to paired blood and CSF samples in neuro-COVID patients (n=23) and HCs (n=21), and the peptides shown in panel b. **e,f** Sequence similarity between **e** the main epitopes of TRIM63 and CCDC63 to the SARS-CoV-2 Spike fusion peptide, and **f** between the main epitopes of TPO and NPC1 and the C-terminal domain of Spike S1.

### Autoantibodies against the main epitopes are found in blood and cerebrospinal fluid from independent cohorts with neuro-COVID but not in pre-pandemic samples

To validate our findings, we analyzed an independent cohort including pre-pandemic healthy controls (HCs, n=29) and neuro-COVID patients (n=25) for the presence of autoantibodies against the detected main epitopes. Cohort demographics are presented in Table S4. Compared with levels in pre-pandemic HCs, the levels of autoantibodies against the main epitopes were significantly elevated in the individuals with the corresponding new-onset autoantibody. In addition, anti-SNURF|50-64, anti-NPC1|566-580, anti-TRIM63|234-247, and anti-TRIM63|236-249 IgG levels were significantly increased in neuro-COVID patients compared to pre-pandemic HCs (Fig. 5b). Notably, the only autoantibody for which the pre-pandemic HCs had levels above background was the previously described autoantibody anti-TPO IgG.

As we were specifically interested in the possible neurological pathology of autoantibodies, we asked whether epitope-directed autoantibodies could also be found in cerebrospinal fluid (CSF) of individuals with COVID-19. In the validation cohorts, paired CSF and blood samples were available for 23 neuro-COVID patients and 21 pre-pandemic HCs. As seen in Figure 5d, epitope-directed autoantibody signals correlated in CSF and blood of both validation cohorts (neuro-COVID: ρ = 0.82, p < 2.2×10^-16^; HCs: ρ = 0.56, p = 9.5×10^-13^).

### The muscle proteins TRIM63 and CCDC63 align with the SARS-CoV-2 fusion peptide

One possible explanation for the emergence of new-onset autoantibodies is molecular mimicry between viral and human proteins. Therefore, we examined any amino acid sequence similarity of the main epitopes and the SARS-CoV-2 Spike glycoprotein^32^ (UniProt accession P0DTC2) using BLAST^33^. We found sequence similarities between the Spike glycoprotein sequence 816-SFIEDLLFNK-825 and TRIM63|234-247 (E=0.022), TRIM63|236-249 (E=0.022), and CCDC63|175-189 (E=0.012) (Figure 5e). This sequence is proximal to the S2’ cleavage site of Spike protein fusion peptide. Furthermore, we found an alignment of NPC1|566-580 and the Spike S1 C-terminal domain sequence 656-VNNSY-660 (E=0.60), which is exposed at the surface of S1. In addition, NPC1|566-580 and TPO|918-932 share five residues (Figure 5f).

## Discussion

In the present work, we have characterized the autoantibody response emerging with COVID-19 using proteome-wide autoantibody screening in longitudinal and independent cohorts. As the scope of our study was a proteome-wide analysis of autoantibody repertoires, we searched for autoantibodies towards intracellular as well as extracellular and secreted antigens. Although the pathogenic mechanisms of antibodies towards intracellular antigens remain unclear, they are frequently observed in screening studies and can be of established clinical importance^34^.

While previous studies have indicated existence of new-onset autoantibodies in COVID-19^14,15^, we have systematically charted the temporal dynamics of the emerging self-directed humoral response in COVID-19 and showed that 60% of new-onset autoantibodies remained elevated for at least 12 months after infection. Our study shows that there is large diversity and interindividual heterogeneity of new-onset autoantibodies in COVID-19, corroborating previous findings in cross-sectional autoantibody studies in health^13^ and disease^31,35^, including COVID-19^11^. Nevertheless, the high prevalence and persistence of new-onset autoantibodies in COVID-19 indicates that dysregulated humoral immunity is a marked feature of acute and post-acute COVID-19, as in many viral infections. The detected new-onset autoantibodies target a wide range of antigens across the proteome, illustrating the breadth of the autoantibody response.

The dynamics of detected new-onset autoantibodies followed three distinct patterns: stable, transient, and delayed onset. The two acute onset types reflect different autoantibody persistence, while delayed onset may reflect other parameters. As 31% (49 of 159) of delayed new-onset autoantibodies emerged in individuals who were vaccinated between seroconversion and the subsequent visit, the onset of these autoantibodies may be vaccine associated. The processes behind the remaining 69% are not clear. We speculate that a short time between infection and blood sampling could prevent immediate detection of new-onset autoantibodies despite detection of seroconversion, possibly due to the high sensitivity of the serological assay^26^. Alternatively, delayed autoantibody onset could reflect autoantibody emergence in late affinity maturation. Further work is needed to shed light on the mechanisms underlying emergence of new-onset autoantibodies.

As anti-IFN IgG is implied in severe COVID-19, we specifically characterized the new-onset anti-IFN IgG response in our study. Considering all IFN subtypes, we found new-onset anti-IFN IgG in 3% of the baseline seronegative cohort, mainly consisting of anti-type I IFN IgG. These results are corroborated by previous studies indicating that anti-IFN IgG in COVID-19 are mainly directed against type I IFNs and that these autoantibodies typically exist prior to COVID-19^1^.

Since neurological symptoms after COVID-19 are commonly occurring and often debilitating, we made directed efforts in understanding this group of symptoms post-COVID-19. We found three new-onset autoantibodies connected to increased severity of neuropsychiatric symptoms post-COVID-19: anti-CALU, MYO16, and SNURF IgG. CALU is a membrane-bound or secreted calcium-binding protein mainly expressed in heart and skeletal muscle. MYO16 is a cytoplasmic unconventional myosin with enhanced brain expression, and may be involved in the extension of neuronal membrane processes^36^. SNURF is a small (71 aa) nuclear protein of unknown function, primarily expressed in brain and muscle tissues and cardiomyocytes. In addition, we found associations of 11 self-assessed non-neurological symptoms post-COVID-19 to 8 of the most prevalent new-onset autoantibodies. However, these associations were tentative, with only the association of anti-PCYT1B IgG and impaired hearing remaining significant after FDR correction.

Our epitope mapping identified the main epitopes of five autoantigens. These epitopes showed a marked increase in reactivity after infection. Among these, the main epitopes of TRIM63 and CCDC63 displayed a sequence alignment to the fusion peptide of the SARS-CoV-2 spike glycoprotein. While not sufficient in isolation, the high post-infectious reactivity to these autoantigens combined with their sequence alignment to a viral protein are together indicative of molecular mimicry. The fusion peptide is crucial for viral entry into the host cell and is highly conserved across the family of coronaviruses. Furthermore, it is accessible to antibody binding in the post fusion state, after engaging the ACE2 receptor^37^. Concordantly, two studies have independently found that human antibodies that broadly neutralize coronaviruses are targeting the fusion peptide^37,38^. This raises the possibility that the observed mimicry also might be found in infections with other coronaviruses. To our knowledge, however, these antigens have not previously been investigated in this context.

With the independent cohort, we validated the presence of anti-TRIM63|234-247, and anti-TRIM63|236-249 IgG in patients with neuro-COVID but not in pre-pandemic HCs. In the discovery cohort, anti-TRIM63 IgG prevalence was increased in hospitalized COVID-19 patients. In addition, we observed a moderate association of anti-CCDC63 IgG and self-reported muscle and joint pain. Taken together, there is evidence for molecular mimicry between the immunologically important Spike protein fusion peptide and the muscle proteins TRIM63 and CCDC63 with early indications of connections to muscular symptoms post-COVID-19 and COVID-19 severity.

Despite not showing any sequence similarity with SARS-CoV-2, anti-SNURF IgG emerged in 9% of HCW (n=40) and 20% of hospitalized COVID-19 patients (n=9). Furthermore, antibodies against the main epitope of SNURF (aa 50-64) were validated in the blood and CSF of patients with neuro-COVID (compared to pre-pandemic HCs). Our detected correlation of autoantibody levels in blood and CSF corroborates findings of blood-brain barrier disruption in patients with cognitive impairment post-COVID-19^39^. It is worth noting that SNURF, like the other two most prevalent autoantibody targets TRIM63 and CCDC63, is expressed in heart and skeletal muscle. In addition, SNURF is expressed in several other tissues including the brain, corroborating the association of anti-SNURF IgG and neuropsychiatric symptoms post-COVID-19. Like CCDC63 and many classical autoantigens, SNURF is located to the nucleus, indicating that epitope spreading may be the source of anti-SNURF IgG^40^. Furthermore, anti-SNURF IgG was by far the most common autoantibody increasing at vaccination. While the data did not sufficiently support an interaction effect of previous new-onset anti-SNURF IgG and vaccine type on anti-SNURF IgG increase at vaccination, this could be due to small sample sizes. However, a recent study reported that autoantibodies remained stable after vaccination with SARS-CoV-2 mRNA vaccines and were not elevated in patients with vaccine-associated myocarditis^15^.

Previous longitudinal studies have reported pre-existing but not new-onset autoantibodies to the clinically important autoantigen TPO in COVID-19 patients^14^. In contrast, we found new-onset anti-TPO IgG in 4% (n=22) of the longitudinal cohort, with a higher prevalence in hospitalized COVID-19 patients than in HCW with mild to moderate disease. In addition, we showed that new-onset anti-TPO IgG co-occurred with new-onset anti-NPC1 IgG in 10 of 22 cases, and anti-NPC1 IgG appeared without anti-TPO IgG in only four individuals. Subsequent epitope mapping and sequence alignment revealed that the main epitopes of TPO and NPC1, which display elevated autoantibody levels after infection, have a sequence similarity containing five identical residues. In addition, the N-terminal residues of the main epitope of NPC1 align with the amino acid sequence 656-VNNSY-660 at the C-terminal domain of the Spike glycoprotein S1 subunit. Together, this raises the possibility of molecular mimicry of NPC1 and Spike S1, with epitope spreading through molecular linkage yielding anti-TPO IgG^40^. Although anti-TPO IgG benignly occurs in around 10% of the healthy population^41^ our discovery of new-onset antithyroid antibodies emerging with mild to severe COVID-19 is concerning given the reports of Grave’s disease, Hashimoto’s disease, and myalgic encephalomyelitis/chronic fatigue syndrome as complications following COVID-19^20,42^. However, we did not detect any anti-TPO IgG increases at vaccination.

Certainly, our study has limitations. Although powerful, the Proteome-wide arrays do not detect autoantibodies towards epitopes that are not covered by the protein fragments on the arrays, or conformational epitopes. Furthermore, as symptoms post-COVID-19 were obtained by self-assessment, clinical associations are limited and require validation in cohorts where symptoms post-COVID-19 have been assessed by a clinician. Moreover, as our controls were healthy and without other infections, the detected new-onset autoantibodies might also develop after other infections than SARS-CoV-2 infection. In addition, the functional properties of the presented autoantibodies remain tentative, and cross-reactivity should be further evaluated in future work using competitive assays.

In summary, our study shows that new-onset autoantibodies are prevalent and persistent in mild to severe COVID-19. Furthermore, some were found to be associated with neuropsychiatric symptoms post-COVID-19 and could be detected in both plasma and CSF of patients with neuro-COVID. In addition, we demonstrate that anti-TRIM63 and anti-CCDC63 IgG develop in 10% of the study cohort and reveal their main epitopes using epitope mapping. The main epitopes display molecular mimicry with the highly conserved fusion peptide of the SARS-CoV-2 Spike glycoprotein which is essential for viral entry and the target of broadly neutralizing antibodies. Conversely, anti-SNURF IgG is highly prevalent without evidence of molecular mimicry, which may indicate epitope spreading to nuclear antigens. Our work reveals the complexity of the autoantibody repertoire that emerges with COVID-19 and shows its potential impact on the course of acute viral infection and post-viral syndromes. This provides a strong rationale for further exploration of new-onset autoantibody repertoires in other infectious diseases, as well as for continued investigation of the herein presented new-onset autoantibodies.

## Methods

### Study cohorts

The COMMUNITY study is an ongoing longitudinal study which enrolled 2149 HCW and 118 COVID-19 patients admitted to Danderyd Hospital, Stockholm, Sweden, between April and May/June 2020 (HCW/patients)^27–29^. The cohort is followed with blood sampling every four months and we assess anti-SARS-CoV-2 IgG using a multiplex bead array of SARS-CoV-2 proteins^26^. In addition, HCW reported symptoms post-COVID-19 through electronic self-assessment forms at selected visits including visits 3 to 5. The symptoms for self-assessment were anxiety, brain fatigue, impaired concentration, cough, depressed mood, diarrhea, dyspnea, dizziness, fatigue, fever, hair loss, headache, impaired hearing, impaired memory, ageusia, muscle/joint pain, nausea, numbness, anosmia, palpitations, skin disorders, sleep disturbance, and stomach ache. Symptom severity was graded as mild, moderate, or severe. Neuropsychiatric symptoms were defined as reporting one or several of anxiety, brain fatigue, impaired concentration, depressed mood, and impaired memory.

In the present study, we retrospectively considered visits 1 to 5, i.e., May 2020 to September 2021. We selected a subgroup of 478 HCW and 47 hospitalized COVID-19 patients based on previous serological results and reported symptoms post-COVID^27–29^. HCW were selected in two steps. First, we selected HCW who were seronegative for anti-SARS-CoV-2 IgG at the first visit in Apr-May 2020, seroconverted prior to vaccination, and had participated in the visits immediately before and after seroconversion (n=381). Second, we selected HCW who were seropositive at the first visit, had participated in all of the four subsequent visits, and had reported several symptoms post-COVID-19 (n=97). Patients were selected based on participation in all of the first four follow-up visits (n=47). In total, 525 individuals were selected with on average 4.8 samples per individual, yielding 2532 samples for autoantibody analysis. Demographics are presented in Table S1.

Ethical approval for the COMMUNITY study was obtained from the Swedish Ethical Review Authority (dnr 2020–01653 and 2021-04113). All health care workers left written informed consent for study participation. For the patients, oral informed consent was obtained instead of written informed consent due to risk of contagion. In the case of incapacity, informed consent was obtained from patients’ next of kin. Oral informed consent was recorded in each patient’s medical record as well as in a separate file held by the responsible researcher. The use of oral consent was approved by the Swedish Ethical Review Authority.

The pre-pandemic healthy control group was university employees and students who had not received psychiatric care during their lifetime and were recruited in the time frame April 2014 - April 2017 as part of the Uppsala Psychiatric Patient Samples (UPP) cohort. Participants underwent a clinical health examination including blood pressure and body mass index (BMI) and answered questionnaires on socio-demographics, medical history, heredity, and current medication, as well as an interview to evaluate any psychiatric symptoms^43^. Demographics are presented in Table S4.

The neuro-COVID population has been described previously^44,45^. Patients were prospectively included between April 2020 and June 2021. Inclusion was based on a positive PCR for SARS-CoV-2 in upper and/or lower airway samples, and at least one new-onset neurological symptom and presence of anti-SARS-CoV-2 IgG in serum, or typical COVID-19 symptoms in combination with pulmonary ground-glass opacities and consolidations on computed tomography scan of the thorax. Clinical neurological evaluation was performed by an experienced neurologist. CSF sampling was performed as part of the routine evaluation when clinically indicated. Demographics are presented in Table S4.

The collection and analysis of neuro-COVID and healthy control samples was approved by the Swedish Ethical Review Authority (dnr 2017-043 with amendments 2019-00169, 2020-01623, 2020-02719, 2020-05730, 2021-01469, and 2020-01883; dnr 2014/148; and dnr 2022-00526-01). The Declaration of Helsinki and its subsequent revisions were followed.

### SARS-CoV-2 serology

Serological classifications for sample selection were obtained from previous studies of the cohorts^27–29^. For increased resolution in the upper ranges of the data, samples were re-analyzed at a higher dilution (1:5000 vs 1:50) while otherwise following the same procedure^26^.

### Planar arrays

Initial exploration of autoantibody repertoires was performed using two sets of in-house developed protein arrays. The Proteome-wide planar array contains 42 000 protein fragments from the Human Protein Atlas (proteinatlas.org) that represent 18 000 proteins and cover approximately 40% of the amino acid residues of the human proteome^31,46^. The Secretome array contains 1522 full-length secreted or extracellular proteins from 1482 genes representing 58% of the human secretome, i.e., the proteins secreted by human cells^30^. The experimental procedure has been described previously^13^. In brief, plasma samples were pooled within the described groups and diluted 1:100 in before applying them to planar arrays. After incubation and washes, fluorescently labelled secondary detection antibodies were applied to the array for detection of autoantibody binding and detection of array microspots. Readout was performed in a microarray scanner, where the red channel readout corresponds to autoantibodies and green channel readout to array microspots. Grid alignment and data acquisition from the scanned images was performed using GenePix Pro 5.1.

### Bead arrays

Bead arrays were used for investigation of new-onset autoantibodies in the COMMUNITY cohort and for epitope mapping in the COMMUNITY and validation cohorts. Bead array construction and assays were performed as previously described^13,47^. For exploration of new-onset autoantibodies, the coupled antigens had been selected from planar arrays as described below. For epitope mapping, the coupled antigens were 93 custom synthesized biotinylated peptides of 14 to 15 amino acid residues (GenScript Biotech). Assay readout was performed using Luminex FLEXMAP 3D® instruments (Luminex Corp.) and responses recorded as median fluorescent intensity, in arbitrary units (MFI AU).

### Analysis of planar array data

Planar array data was processed as previously described^31^.

Proteome-wide array data was analyzed per subarray. Spots that were flagged in image acquisition, that were smaller than 30 pixels, or that had a green channel signal lower than 4 SD above the mean local background were removed. Duplicate spots were deduplicated by selecting the spot with highest signal in the green channel. Finally, red channel data were Z-scored, and antigens with Z ≥ 12 were classified as reactive.

Secretome array data was analyzed per subarray. Microarray spots that were flagged in image acquisition, that were smaller than 40 pixels, or that had a green channel signal at or below the local background level were removed. Duplicate spots were filtered if their CV exceeded 50, and remaining spot pairs were deduplicated by taking the mean. Finally, local background was subtracted from red channel data and resulting values were Z-scored. Antigens with Z ≥ 8 were classified as reactive.

Z-scored planar array data were used for selection of antigens for further investigation in the full HCW and hospitalized patient cohorts. First, antigens reactive in single samples on single arrays were selected. Second, lowered selection thresholds and detection in multiple samples were used to diversify the selection. The following selection criteria were used: protein fragments meeting Z ≥ 8 in multiple samples; full-length proteins meeting Z ≥ 4 in multiple samples; protein fragments and full-length proteins meeting Z ≥ 4 in single or multiple samples on both array types. Third, antigens from the literature and in-house studies were selected: protein fragments noted in both the literature and in-house studies; protein fragments meeting Z ≥ 8 and noted in either the literature or in-house studies; full-length proteins noted in multiple publications; full-length proteins meeting Z ≥ 2 and noted in the literature; handpicked protein fragments and full-length proteins noted in either the literature or in-house studies. Selected antigens and their matching selection criteria are listed in Table S2.

### Classification of new-onset autoantibodies in individuals with a seronegative baseline sample

New-onset autoantibodies were classified by applying Partitioning Around Medoids (PAM) clustering^48^ to the bead array data of baseline seronegative individuals.

The log_2_ FC of each autoantibody trajectory was computed relative to the most recent seronegative sample. Individuals with autoantibody data at seroconversion and the samplings immediately before and after seroconversion were considered for PAM clustering (n=374). Principal component analysis (PCA) was conducted on these data to identify and exclude outlying individuals with an increase in most autoantibody trajectories at seroconversion (n=5, Fig. S7a). Outliers were defined as PC1≥mean+3×SD of PC1 (PC1≥8.85). To reduce the running time of the model, trajectories that never exceed background MFI levels were pre-classified as not new-onset. In each antigen class (protein fragments and full-length proteins), background MFI level was defined as robust Z score = 3 in the seronegative samples from September 2020 and January 2021 from the 40 individuals that seroconverted in May 2021 (MFI_fragment_≥73, MFI_full-length_≥322; n filtered trajectories = 619894/910041 (68%)).

The PAM clustering was performed with the custom cosine × euclidean distance metric. To evaluate the parameter k (number of clusters), the PAM clustering was run with k ranging from 1 to 20, each with 10 random starts of the clustering algorithm. Using the silhouette method, the optimal cluster number was determined to be 7. Based on a median log_2_ FC ≥ 2, cluster 5, 6, and 7 were classified as new-onset.

### Classification of new-onset autoantibodies in individuals without any seronegative baseline sample

New-onset autoantibodies were assessed in individuals without seronegative baseline samples using Multinomial Linear Regression (MNL)^49,50^.

The MNL model was built using classifications and autoantibody trajectories of the 22 prevalent new-onset autoantibodies in the baseline seronegative individuals that had follow-up samples at 4 and 8 months after seroconversion. The MNL model specification was 𝑁𝑒𝑤– 𝑜𝑛𝑠𝑒𝑡 𝑡𝑦𝑝𝑒 ∼ log_10_ 𝑀𝐹𝐼_𝑡(0)_ + 𝐹𝐶_𝑡(4)_ + 𝐹𝐶_𝑡(8)_, where *t* is months from seroconversion, and *FC* is fold change relative to time of seroconversion (*t(0)*). The trajectories were split into training (0.8) and testing (0.2) sets using randomized stratified sampling on the trajectory outcome (target antigen and simplified new-onset classification; acute, delayed, or not new-onset autoantibody). The MNL model was trained using repeated stratified K-fold cross-validation (repeats = 10, K = 5, stratification on trajectory outcome). Grid search was used to optimize the penalizing decay parameter, which was set to 0 based on model accuracy.

Individuals that did not have seronegative baseline samples, but that did have autoantibody data at seroconversion and the two samplings immediately after seroconversion, were considered for MNL classification (n=135). PCA was used to exclude outlying individuals with an increase in most autoantibody trajectories in the considered timepoints (n=2, Fig. S7b). Outliers were defined as PC1≥mean+3×SD of PC1 (PC1≥2.1). Trajectories of the 22 prevalent new-onset autoantibodies in the remaining 133 individuals were classified using the MNL model (n trajectories = 2926).

### Annotation of antigen location

Antigen location was determined based on Cellular Component Gene Ontology (GO) terms for each corresponding gene^51,52^. The GO terms were simplified using the Generic GO subset^53^ and further subdivided into broad location categories as detailed in Table S5. For antigens with multiple locations, the annotation was selected in the following order: Extracellular > Plasma membrane > Nuclear > Intracellular. Antigens may lack annotation of location if no parent terms are included in the GO subset.

### Association of new-onset autoantibodies with symptoms post-COVID-19

Association of new-onset autoantibodies with symptoms post-COVID-19 was performed using data from HCW displaying one or more of the 22 prevalent new-onset autoantibodies, or none (n=403). Among these, neuropsychiatric symptoms were defined as reporting anxiety, brain fatigue, difficulties concentrating, depressive disorders, or impaired memory that lasted for at least 2 months after COVID-19 (n=384). The highest reported severity was used (severe=20 (5%); moderate=58 (15%); mild=23 (6%); no neuropsychiatric sx=283 (74%)). For other symptoms post-COVID-19, reported mild symptoms were excluded (Table S6).

### Identification of main epitopes

The peptide bead array was used for epitope mapping. Data were acquired from 142 baseline samples and 150 samples after autoantibody onset in the 142 individuals that had one or more new-onset autoantibodies whose antigens were represented on the peptide bead array. (Eight individuals had both acute and delayed new-onset autoantibodies, increasing the number of samples after autoantibody onset.) The FC was computed relative to baseline, both for beads with coupled peptides as well as the negative control bead with coupled biotin. To adjust for individual background levels, the FC of biotin was subtracted from the FC of the peptides, and the median was set to 1. Any resulting negative values (n=2) were imputed with the smallest positive value present (0.07).

Main epitopes were defined in individuals having the corresponding new-onset autoantibody by taking the mean FC of each peptide. Peptides where the mean exceeded the background FC cutoff were defined as main epitopes that were common across individuals. The background FC cutoff was defined as mean+5×SD of the FC of the negative biotin control (FC≥2.52).

### Sequence alignment

Sequence alignment was performed using NCBI BLASTP adjusted for short input sequences: E < 20 000, word size ≥ 3, PAM30 substitution matrix, gap costs 7/2, no compositional adjustment, and no filters or masks.

### Statistical analysis

Correlations were performed using Spearman correlation. Group differences of continuous variables were investigated using the Mann-Whitney U test. Binary variables were investigated using logistic regression, categorical variables using multinomial logistic regression, and ordinal variables using proportional odds logistic regression. Regression models used correction for age and sex in group comparisons. False discovery rate correction was performed using the Benjamini-Hochberg procedure and resulting values are reported as q-values.

All data analysis and statistical analysis was performed in RStudio with R 4.1.2 and 4.2.1 and R packages tidyverse, lubridate, rlang, scales, knitr, pander, httr2, readxl, rstatix, proxy, cluster, caret, MLmetrics, MASS, broom, Omixer, ggpubr, ragg, patchwork, cowplot, GGally, ggsignif, ggfortify, ggdist, ggbeeswarm, ggrepel, ComplexHeatmap, and circlize.

## Data Availability

All data produced in the present study are available upon reasonable request to the authors and approval by the relevant ethical boards. Data cannot be shared freely as it contains sensitive personal data covered by the GDPR.

## Acknowledgements

This research was funded by the Swedish Research Council (registration number 2022-03063). The collection of the neuro-COVID cohort was supported by grants within SciLife-U COVID19, KAW-KTH SciLifeLab, and SLS Covid19. The authors thank Eva Kumlien and Johan Virhammar for providing control samples; the KTH node of Protein Production Sweden (PPS), a national research infrastructure funded by the Swedish Research Council, for providing antigens; Eni Andersson, Ceke Hellström, and Jennie Olofsson for assistance with experimental and computational work; Sára Mravinacová, Sofia Bergström, and Jochen Schwenk for fruitful discussions; and all study participants for their participation.

## Author contributions

A.J.F., P.N., A.M., Se.H., and C.T. conceptualized and designed the experiments and study. A.J.F., P.N., A.M., and E.P. interpreted the data and wrote the manuscript. A.J.F. and L.S. performed experimental work and A.J.F. analyzed and visualized the data. So.H., H.T., R.S., and P.N. produced the antigens and developed the arrays. C.T. and Se.H. collected and curated the COMMUNITY cohort. J.L.C., A.R., and E.R. provided the validation cohorts. The manuscript was edited and approved by all authors.

