## Supplementary material for "Prevalent and persistent new-onset autoantibodies in mild to severe COVID-19"

August Jernbom Falk^1^*, Lovisa Skoglund^1^, Elisa Pin^1^, Ronald Sjöberg^1^, Hanna Tegel^2^, Sophia Hober^2^, Elham Rostami^3,4^, Annica Rasmusson^5^, Janet L. Cunningham^5^, Sebastian Havervall^6^, Charlotte Thålin^6^, Anna Månberg^1^, Peter Nilsson^1^

^1^ Division of Affinity Proteomics, Department of Protein Science, SciLifeLab, KTH Royal Institute of Technology, Stockholm, Sweden

^2^ Division of Protein Technology, Department of Protein Science, KTH Royal Institute of Technology, Stockholm, Sweden

^3^ Section of Neurosurgery, Department of Medical Sciences, Uppsala University Hospital, Uppsala, Sweden

^4^ Department of Neuroscience, Karolinska Institutet, Stockholm, Sweden

^5^ Department of Medical Sciences, Psychiatry, Uppsala University, Uppsala, Sweden

^6^ Department of Clinical Sciences, Karolinska Institutet, Danderyd Hospital, Stockholm, Sweden

* Corresponding author

Table S1 | Cohort demographics.

|  | **HCW** | **Hosp. patients** |
| --- | --- | --- |
| **N** | 478 | 47 |
| **N samples [mean (SD)]** | 4.8 (0.5) | 5 (0) |
| **Age [years, mean (SD)]** | 45 (11) | 57 (13) |
| **Sex [F\|M, N (%)]** | 416 (87%) \| 62 (13%) | 16 (34%) \| 31 (66%) |
| **Seroconversion [N (%)]** |  |  |
| **May 2020** | 96 (20%) | 40 (85%) |
| **Sept 2020** | 109 (23%) | 7 (15%) |
| **Jan 2021** | 233 (49%) | 0 (0%) |
| **May 2021** | 40 (8%) | 0 (0%) |
| **Neuropsychiatric symptoms post-COVID-19 [N (%)]** |  |  |
| **Mild** | 29 (6%) |  |
| **Moderate** | 83 (17%) |  |
| **Severe** | 25 (5%) |  |
| **None** | 341 (71%) |  |
| **Any symptoms  post-COVID-19 [N (%)]** |  |  |
| **No** | 184 (38%) |  |
| **Yes** | 294 (62%) |  |

Table S2 | Protein antigens for analysis of the new-onset autoantibody repertoire.

Table presented as a separate excel file.

Table S3 | Peptide antigens for eptitope mapping.

Table presented as a separate excel file.

Table S4 | Validation cohort demographics.

|  | **Neuro-COVID** | **Pre-pandemic HC** |
| --- | --- | --- |
| **Sera (N)** | 25 | 29 |
| **CSF (N)** | 21 | 23 |
| **Age [years, mean (SD)]** | 62 (16) | 24 (6) |
| **Sex [F\|M, N (%)]** | 8 (32%) \| 16 (64%) | 26 (90%) \| 3 (10%) |
| **Age and sex not reported (N)** | 1 | 0 |

Table S5 | Location categories of the Generic GO term subset.

| **GO ID** | **Name** | **Annotation** |
| --- | --- | --- |
| GO:0005576 | extracellular region | Extracellular |
| GO:0005615 | extracellular space | Extracellular |
| GO:0005929 | cilium | Extracellular |
| GO:0030312 | external encapsulating structure | Extracellular |
| GO:0031012 | extracellular matrix | Extracellular |
| GO:0005618 | cell wall | Plasma membrane |
| GO:0005886 | plasma membrane | Plasma membrane |
| GO:0000228 | nuclear chromosome | Nuclear |
| GO:0005634 | nucleus | Nuclear |
| GO:0005635 | nuclear envelope | Nuclear |
| GO:0005654 | nucleoplasm | Nuclear |
| GO:0005694 | chromosome | Nuclear |
| GO:0005730 | nucleolus | Nuclear |
| GO:0005739 | mitochondrion | Intracellular |
| GO:0005764 | lysosome | Intracellular |
| GO:0005768 | endosome | Intracellular |
| GO:0005773 | vacuole | Intracellular |
| GO:0005777 | peroxisome | Intracellular |
| GO:0005783 | endoplasmic reticulum | Intracellular |
| GO:0005794 | Golgi apparatus | Intracellular |
| GO:0005811 | lipid droplet | Intracellular |
| GO:0005815 | microtubule organizing center | Intracellular |
| GO:0005829 | cytosol | Intracellular |
| GO:0005840 | ribosome | Intracellular |
| GO:0005856 | cytoskeleton | Intracellular |
| GO:0009536 | plastid | Intracellular |
| GO:0009579 | thylakoid | Intracellular |
| GO:0031410 | cytoplasmic vesicle | Intracellular |
| GO:0043226 | organelle | Intracellular |

Table S6 | Self-reported symptoms post-COVID-19. Prevalence of self-reported moderate or severe symptoms among HCW. Data pertaining to Figure 3d. Self-reported mild symptoms were excluded.

| **Symptom post-COVID-19** | **Moderate or severe [n(%)]** | **Not present [n(%)]** |
| --- | --- | --- |
| Cough | 46 (11) | 329 (82) |
| Diarrhea | 16 (4) | 374 (93) |
| Dyspnea | 57 (14) | 321 (80) |
| Dizziness | 28 (7) | 356 (88) |
| Fatigue | 144 (36) | 222 (55) |
| Fever | 24 (6) | 372 (92) |
| Hair loss | 37 (9) | 351 (87) |
| Headache | 78 (19) | 301 (75) |
| Impaired hearing | 14 (3) | 384 (95) |
| Ageusia | 117 (29) | 268 (67) |
| Bodily pain | 80 (20) | 305 (76) |
| Nausea | 18 (4) | 369 (92) |
| Numbness | 6 (1) | 379 (94) |
| Anosmia | 130 (32) | 253 (63) |
| Palpitations | 51 (13) | 332 (82) |
| Skin disorders | 16 (4) | 382 (95) |
| Sleep disturbance | 66 (16) | 328 (81) |
| Stomach ache | 10 (2) | 386 (96) |

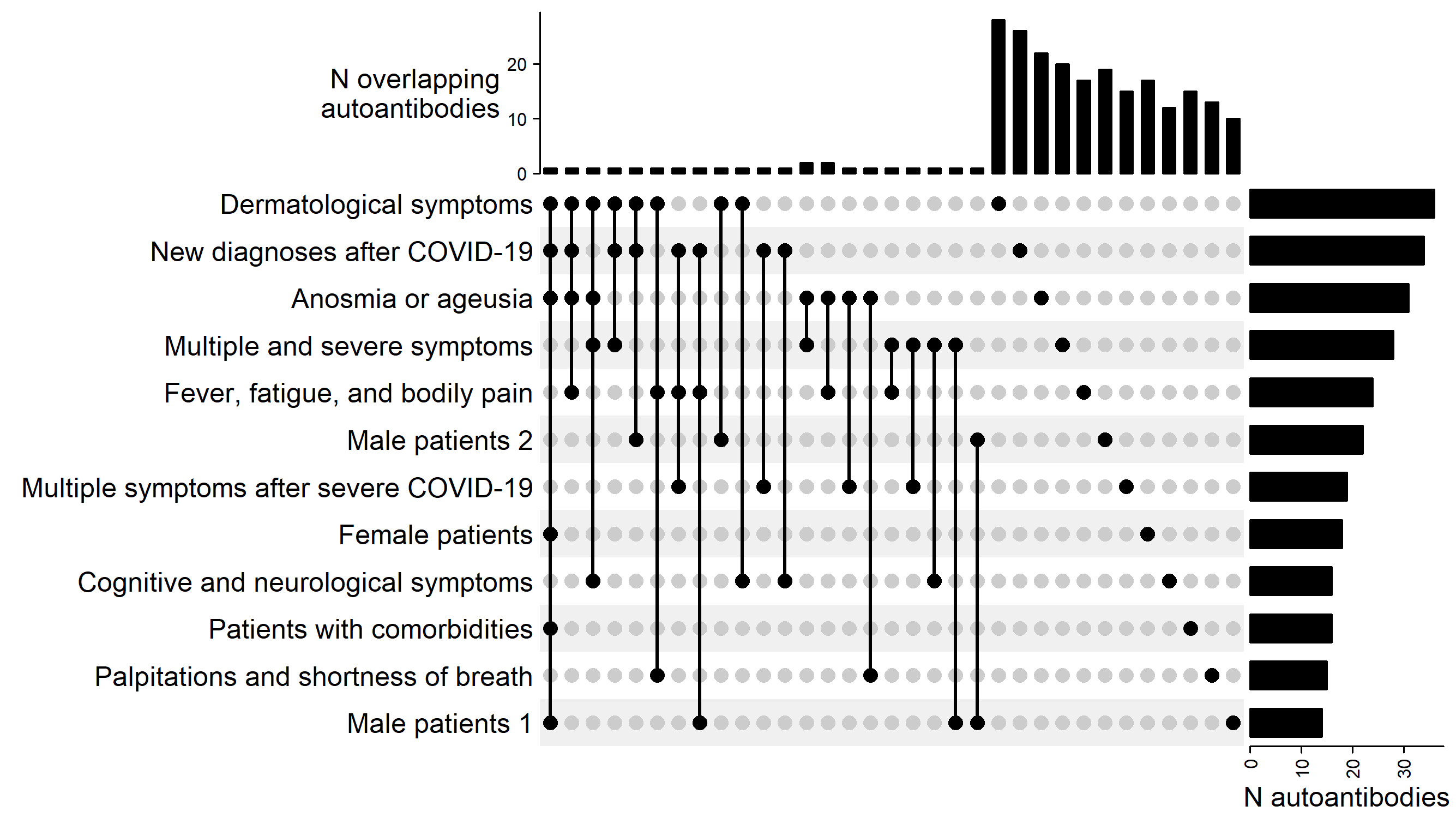

Figure S1 | Planar arrays reveal heterogeneous autoantibody profiles in groups of HCW and hospitalized patients. Row bars depict the number of detected autoantibodies in each group of HCW or hospitalized patients. Column bars depict the intersection of autoantibody profiles, i.e., the number of autoantibodies present in single or multiple groups in accordance with the intersection matrix.

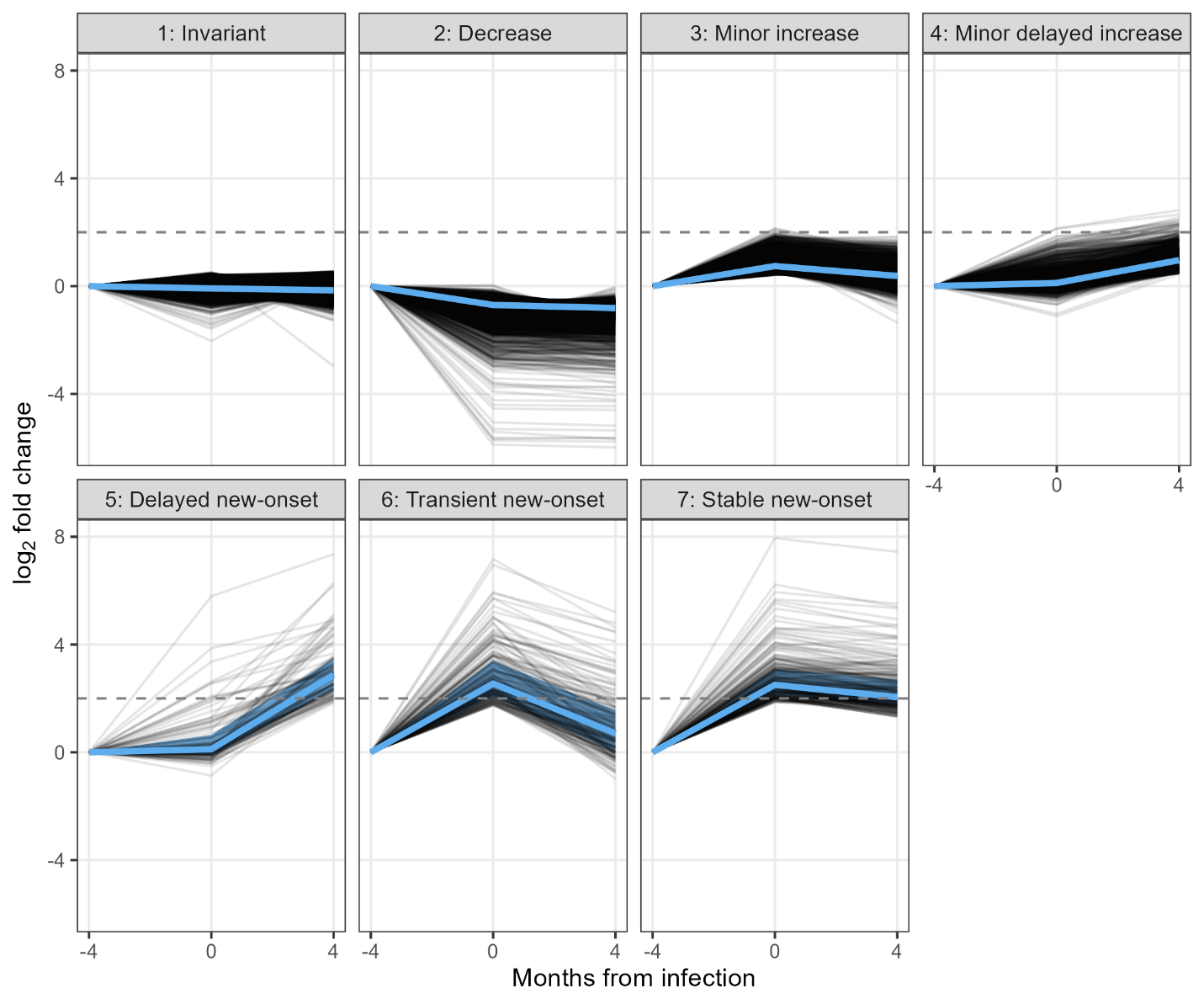

Figure S2 | PAM clustering stratifies autoantibody dynamics.

PAM clustering with the custom cosine × euclidean distance metric revealed 7 clusters of autoantibody dynamics. Black lines depict autoantibody trajectories (of one autoantibody in one individual). Blue lines and shaded areas represent median and quartiles, respectively.

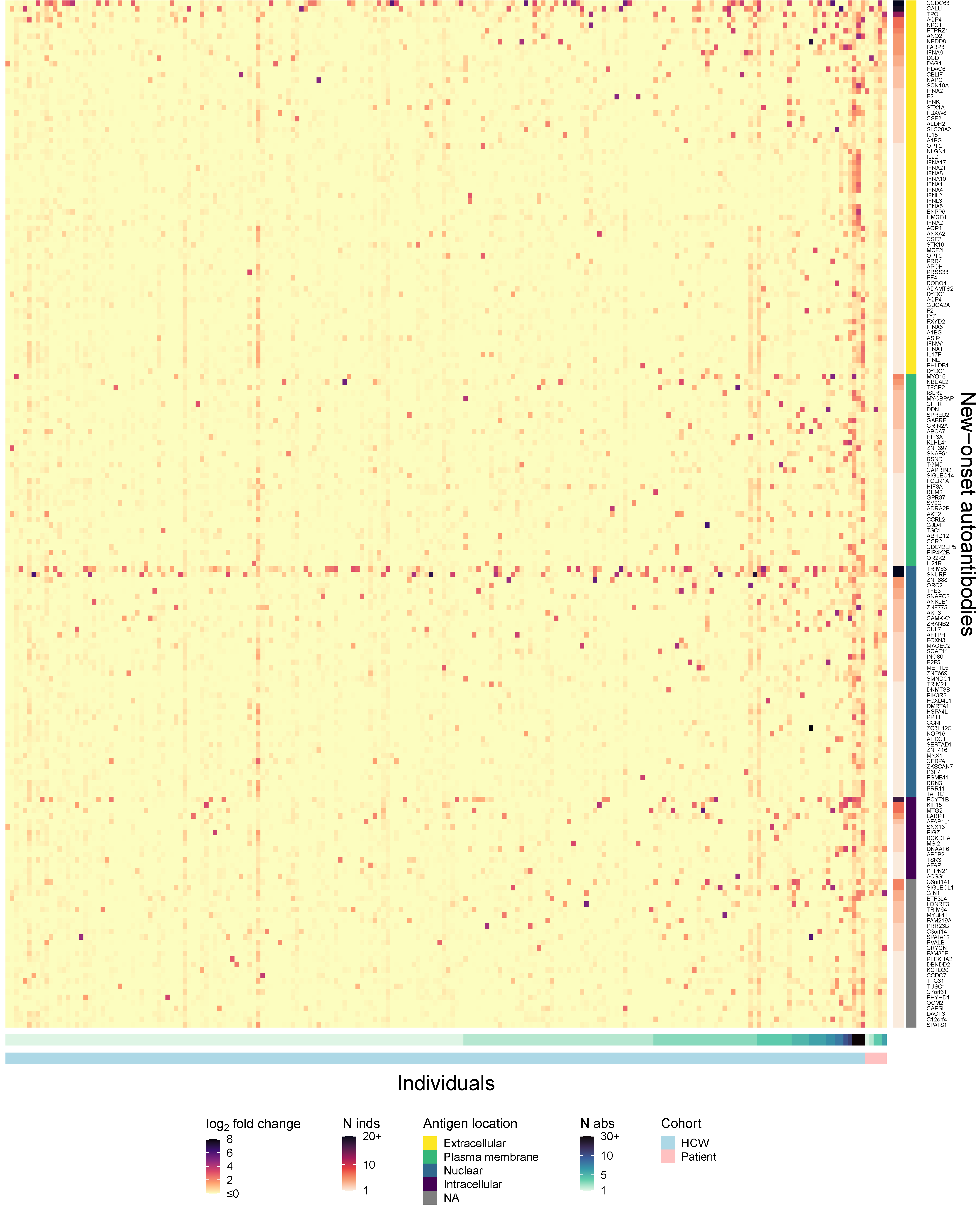

Figure S3 | The autoantibody landscape at seroconversion. Heatmap depicting log_2_ FC of autoantibody signals at seroconversion. Rows depict detected new-onset autoantibodies arranged by antigen location. Antigens may lack annotation due to the GO subset mapping (see Methods). Columns show unique individuals with seronegative baseline samples and at least one detected new-onset autoantibody. Individuals are arranged by increasing number of new-onset autoantibodies and cohort. Cell color represents log_2_ FC of autoantibody signal intensity at infection. Antigen gene names may occur more than once due to multiple protein representations.

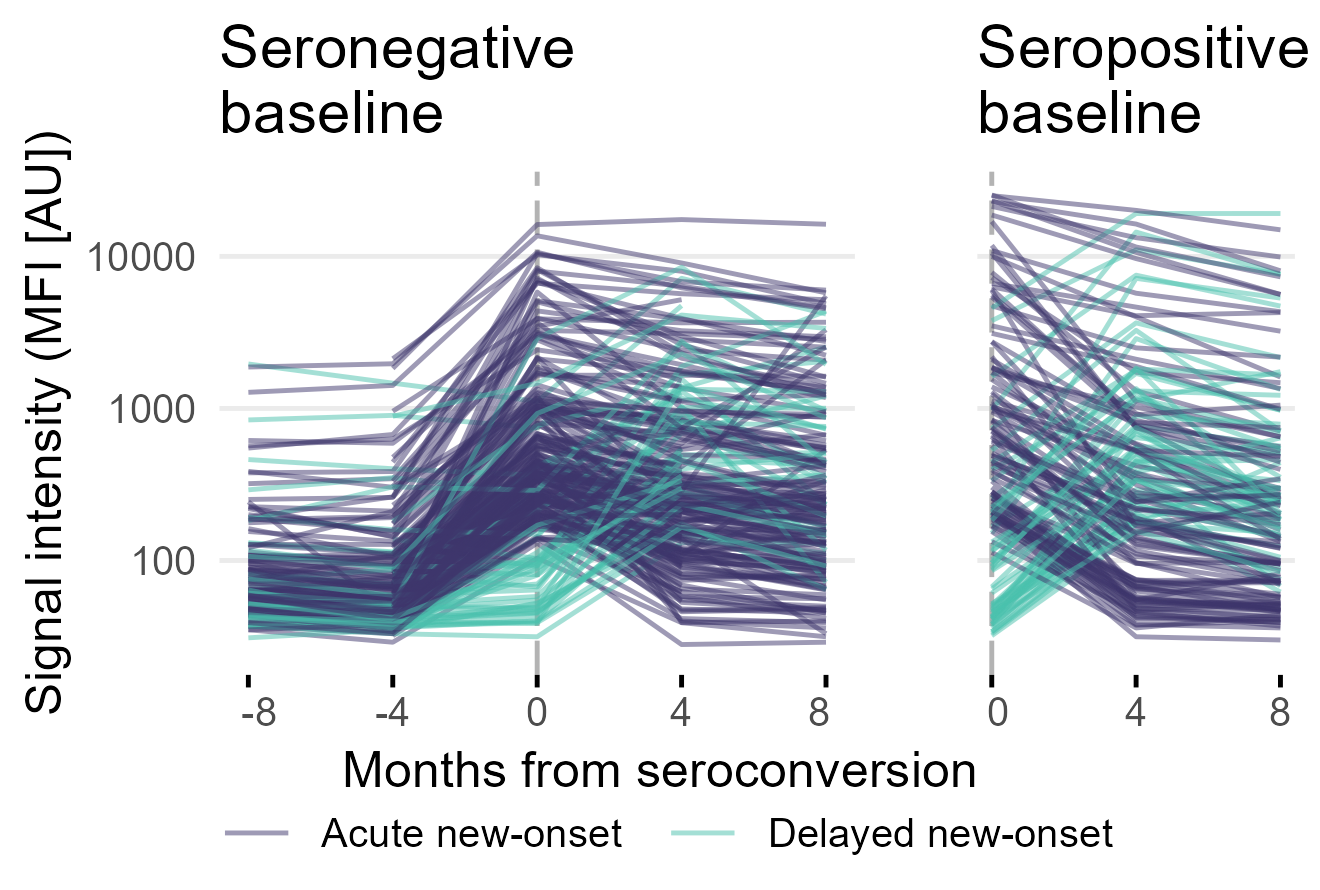

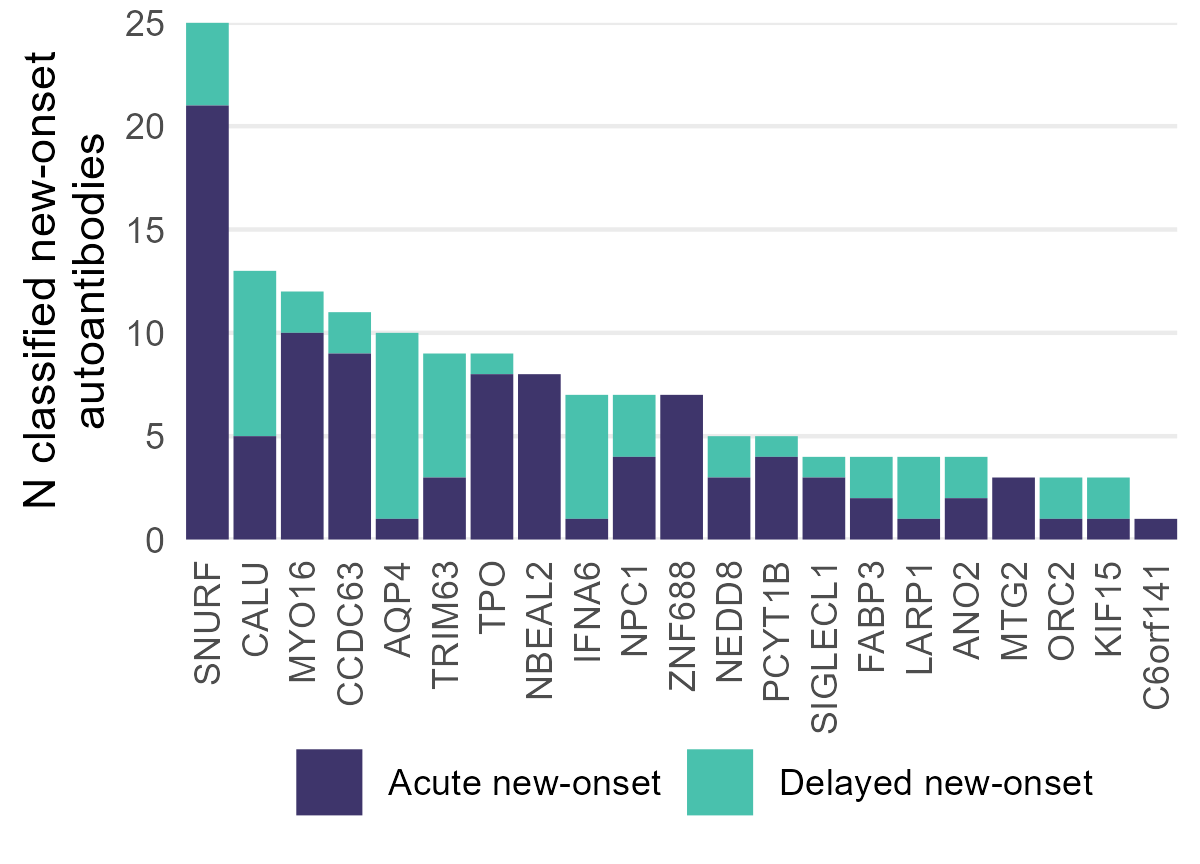

b

a

Figure S4 | Classification of new-onset autoantibodies in individuals without seronegative baseline sample. **a** Trajectories of new-onset autoantibodies in individuals with seronegative (left) and seropositive (right) baseline sample. **b** Count of new-onset autoantibodies in individuals with seropositive baseline sample.

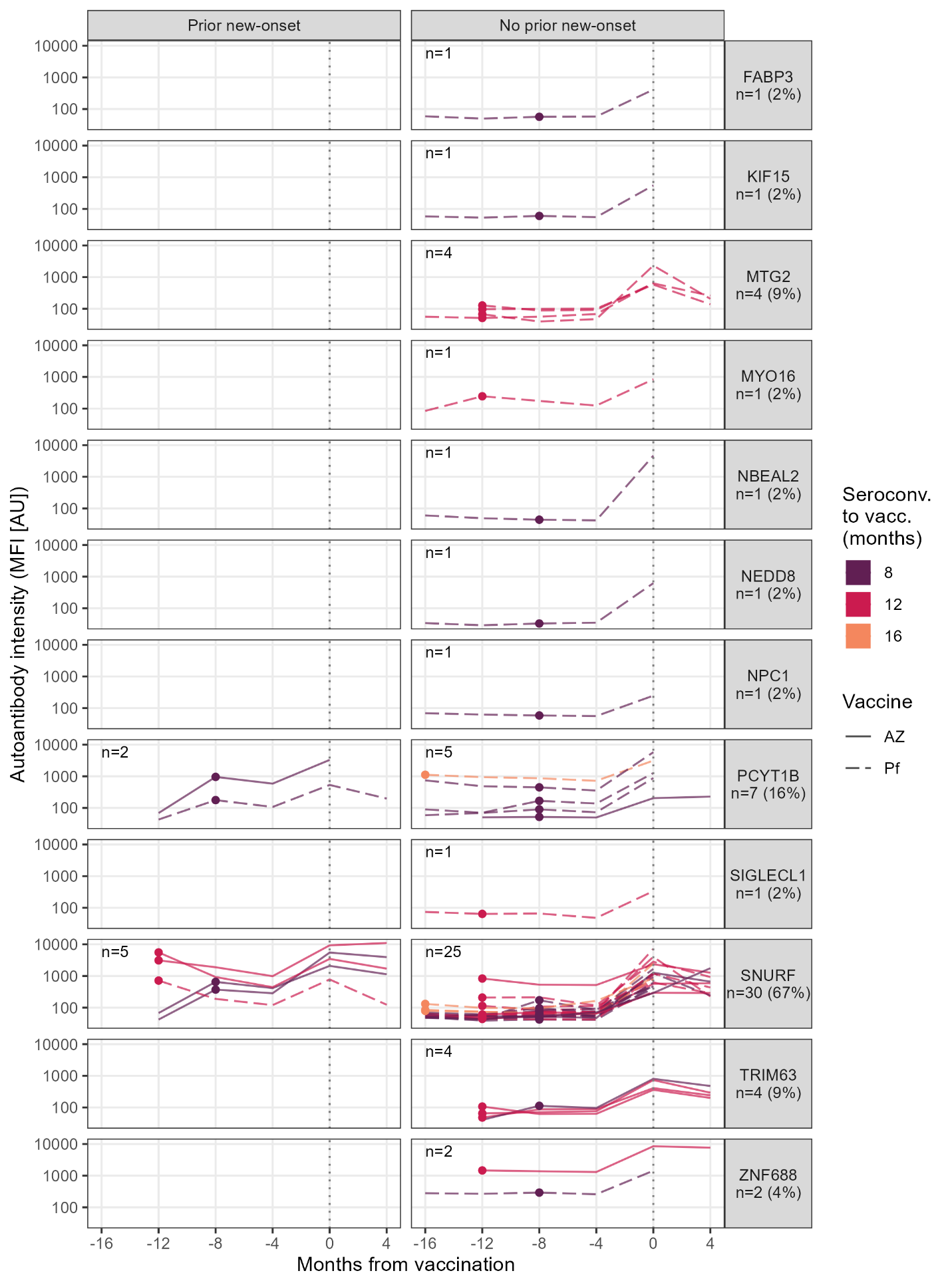

Figure S5 | Increasing levels of new-onset autoantibodies at vaccination. Lines depict individuals with a 4-fold increase of any of the 22 prevalent new-onset autoantibodies at vaccination. Points indicate time of seroconversion. Color indicates months between seroconversion and first vaccination. Individuals are separated across panel columns based on any prior new-onset autoantibody towards the corresponding antigen (panel rows) at infection. Percentages were calculated among all HCW with an autoantibody that increased at vaccination (n=45).

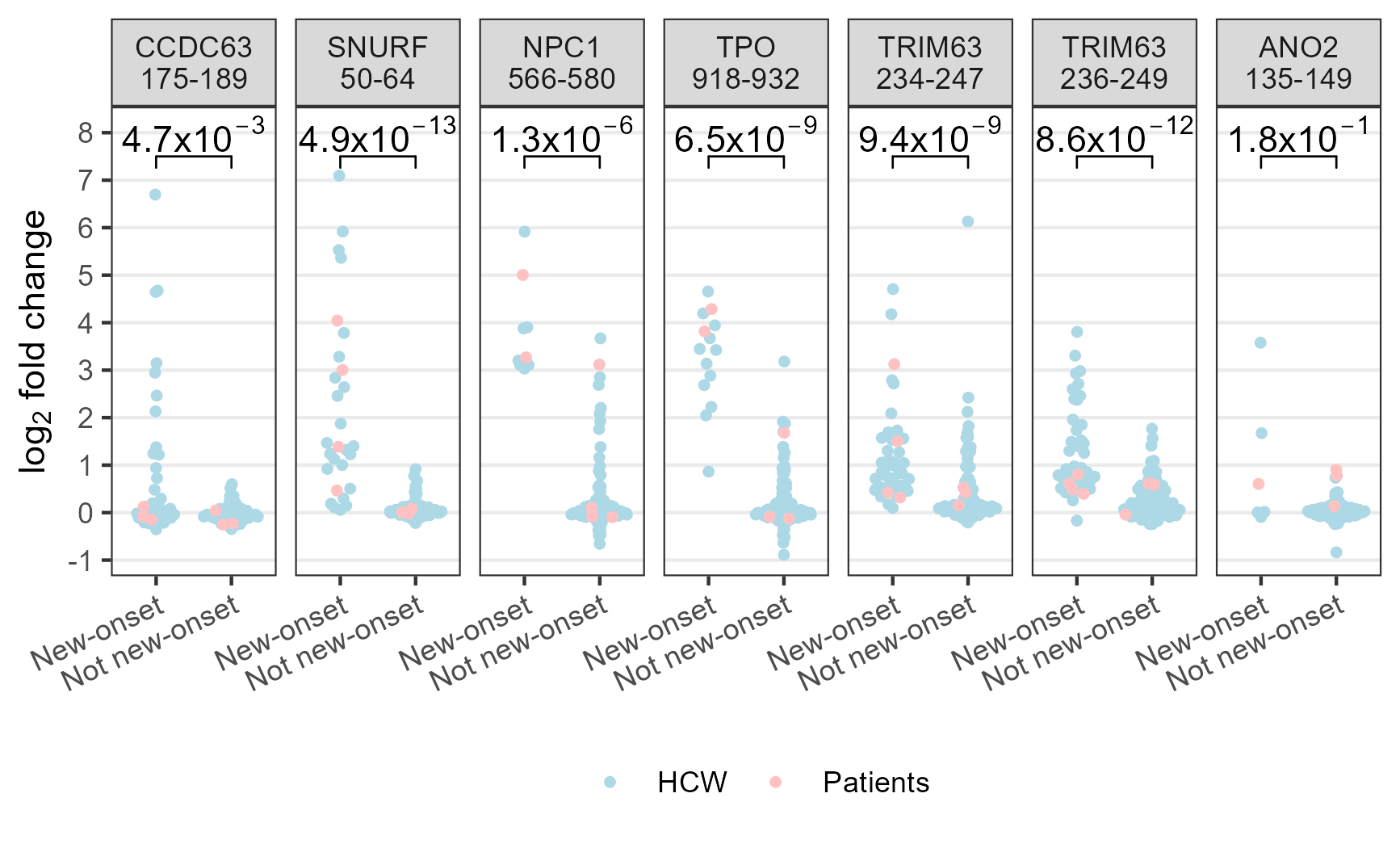

Figure S6 | Autoantibodies against the main epitopes in individuals with vs. without the respective new-onset autoantibody. Points correspond to individuals. Categories on the x-axis indicate the prior classification of individuals’ autoantibody trajectories towards the protein fragment corresponding to the peptide indicated in the panel titles. Levels on the y-axis correspond to the peptide indicated in the panel titles.

a

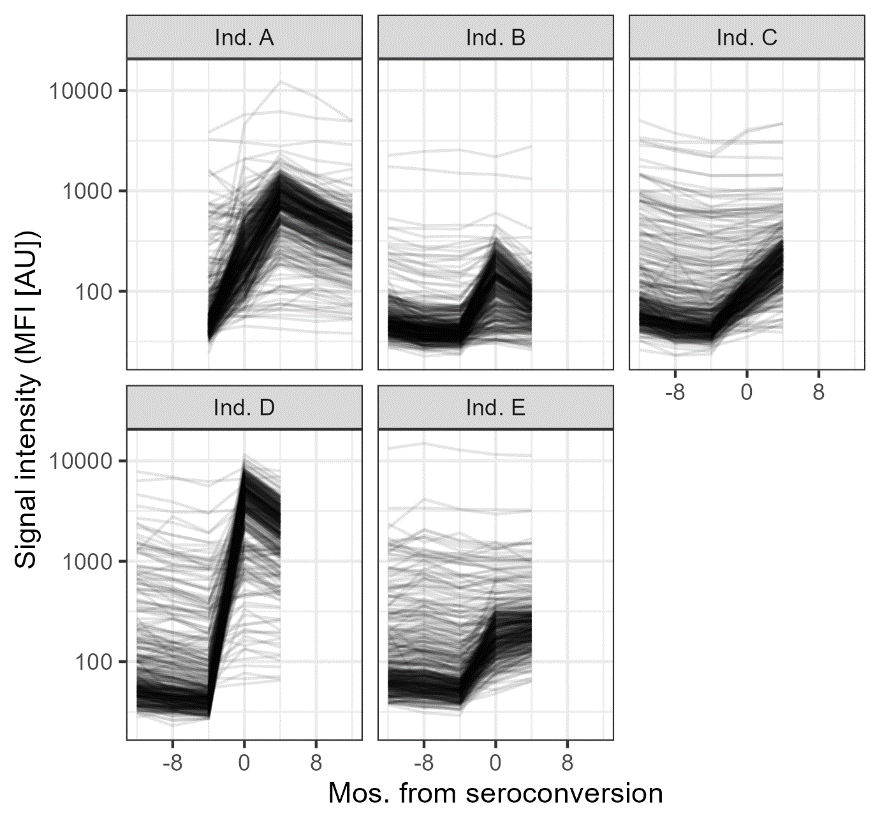

b

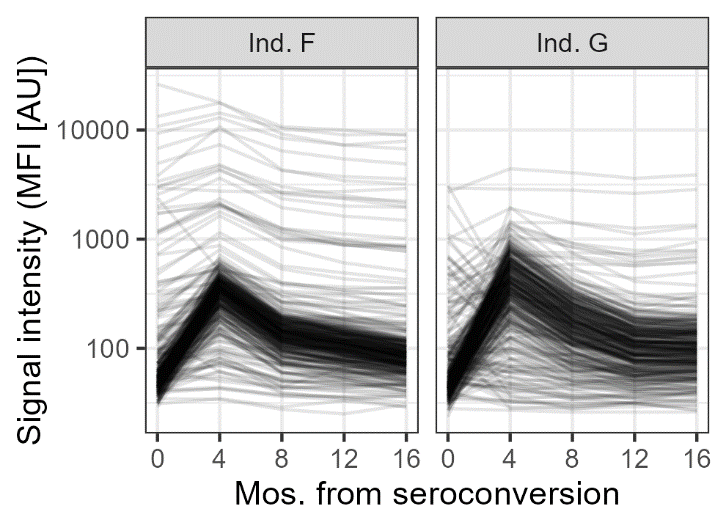

Figure S7 | Individuals with deviating global autoantibody patterns. **a** Autoantibody line plots of the 5 individuals with high fold change at seroconversion across most autoantibodies. These individuals were identified using PCA and excluded from PAM clustering. **b** Autoantibody line plots of the 2 individuals with high fold change across most autoantibodies 4 months after seroconversion. These individuals were identified using PCA and excluded from MNL classification.
